# Mental Health under Pressure: PTSD Risk Among Healthcare Workers in Ekiti State, Nigeria – The Role of Health Beliefs and Experiential Avoidance

**DOI:** 10.1101/2025.08.09.25333324

**Authors:** Dogbahgen Alphonso Yarseah, Ololade Omolayo Ogunsanmi, Viola H. Cheeseman, Elijah Olawale Olaoye, Joyce Olufunke Ogunsanmi, Olu Francis Ibimiluyi

## Abstract

**Background:** Healthcare workers (HCWs) in Ekiti State, Nigeria, faced significant psychological stress during the early phase of the COVID-19 pandemic. However, limited evidence exists on how health behavior, experiential avoidance, and the Health Belief Model (HBM) jointly shape posttraumatic stress disorder (PTSD) symptoms, especially through moderated mediation mechanisms.

**Methods:** A cross-sectional survey of 475 HCWs (171 males, 304 females) was conducted using snowball sampling across two urban local governments. Instruments included the HBM Scale, Health Behavior Questionnaire, Acceptance and Action Questionnaire-II (AAQ-II), and the PTSD Checklist (Civilian Version). Data were analyzed using SPSS v22 and SmartPLS.

**Results:** Health behavior significantly predicted HBM scores (β = 0.156, p = 0.005) but not PTSD symptoms directly (β = 0.097, p = 0.101). Experiential avoidance predicted both HBM (β = –0.143, p = 0.003) and PTSD (β = 0.392, p < 0.001). HBM partially mediated the effects of health behavior (β = –0.022, p = 0.043) and experiential avoidance (β = 0.019, p = 0.045) on PTSD. Conditional indirect effects showed that these mediation pathways were moderated by age (β = 0.028, p = 0.019), years of practice (β = 0.024, p = 0.025), professional designation (β = –0.014, p = 0.036), and experiential avoidance (β = 0.020, p = 0.041). Predictive assessment confirmed strong model performance, especially for PTSD symptoms (Q²predict = 0.222; RMSE = 0.887, MAE = 0.684), compared to HBM (Q²predict = 0.093; RMSE = 0.960, MAE = 0.726).

**Conclusion:** The HBM played a dual role—both buffering and amplifying the effects of behavior and avoidance on PTSD—depending on demographic and psychological context. The model demonstrates both explanatory and predictive value, reinforcing the need for belief-sensitive, tailored interventions to reduce PTSD risk in resource-constrained healthcare environments.

## Background of the Study

The coronavirus disease (COVID-19), one of the most devastating and contagious pandemics in recent history, emerged in December 2019 and rapidly evolved into a global health crisis. By early 2020, the World Health Organization (WHO) had declared COVID-19 a pandemic, with over 108 million infections and 2.38 million deaths reported across 219 countries by February 2021[1]. Although the exact origin remains debated, it is widely believed to have originated in Wuhan, China [2]. This respiratory disease is characterized by symptoms such as fever, fatigue, cough, breathing difficulties, and the loss of smell and taste [3–5].

Although Africa accounted for only 1,472 (1.0%) of global COVID-19 infections and 17 (1.2%) of healthcare worker deaths, the continent had a case fatality rate (CFR) of 1.2%, suggesting a higher proportion of deaths relative to infections compared to regions with more cases [6]. This may reflect persistent mental health challenges in healthcare infrastructure across the continent. Nigeria, Africa’s most populous country with over 223 million people, recorded 266,675 confirmed COVID-19 cases and 3,155 deaths as of May 28, 2023 [7]. This equates to a prevalence rate of approximately 0.12% and a CFR of 1.18% in the general population. While the overall mortality rate was relatively low—around 0.0014%, Nigeria’s CFR was slightly above the global average of 1.1% at that time. Due to its strong travel and economic ties with China, Nigeria was identified early in the pandemic as one of the top 13 countries at high risk of COVID-19 importation [8–9].

In many African contexts, including Nigeria, healthcare workers (HCWs) often relied on delayed or externally sourced information—mainly from Western institutions such as the WHO and CDC—before local understanding of transmission patterns and protective protocols for novel diseases like COVID-19 emerged. This epistemic dependence contributed to uncertainty and diminished professional agency, particularly during the early stages of the outbreak. These conditions fostered experiential avoidance (EA), a psychological tendency to evade distressing internal experiences among frontline HCWs operating in high-risk environments without adequate guidance or consistent public health messaging.

These theoretical concerns became especially visible during the early phase of the pandemic in Ekiti State, where socio-economic disparities and public mistrust in government authorities significantly impeded adherence to preventive health directives. While health officials promoted behaviors such as mask-wearing and social distancing, many citizens, especially those experiencing financial hardship struggled to comply due to the need for daily income. Furthermore, perceptions that government relief efforts were politically biased undermined public trust and cooperation. In this volatile setting, HCWs contended with overwhelming caseloads, insufficient personal protective equipment, and continuous exposure to risk, all while engaging with a skeptical and often resistant population.

These psychosocial stressors contributed not only to emotional exhaustion and burnout but also heightened the risk of developing post-traumatic stress disorder (PTSD). PTSD, as defined by the Diagnostic and Statistical Manual of Mental Disorders, Fourth Edition (DSM-IV) [10], is a psychiatric condition that may arise following exposure to traumatic events and is characterized by symptoms such as intrusive re-experiencing, avoidance, and hyperarousal. Although PTSD symptoms among healthcare workers (HCWs) have been widely documented in global contexts, limited research has explored the psychological mechanisms underlying trauma responses within high-risk African healthcare environments like Nigeria. Specifically, factors such as **experiential avoidance** and **health behaviors** may exert both direct and indirect influences on PTSD outcomes but to our knowledge, no study has examined these causal change mechanism.

Experiential avoidance (EA) refers to the psychological tendency to evade or suppress distressing internal experiences, such as painful emotions, thoughts, or memories [11–13]. Although its effects on PTSD through the Health Belief Model (HBM) remain underexplored in health literature, EA may serve as a short-term coping strategy among frontline healthcare workers (HCWs), particularly in high-stress and unpredictable environments common in many African settings. Over time, however, this avoidance can contribute to maladaptive outcomes, including heightened vulnerability to PTSD. Thus, EA warrants further investigation as a potential risk factor in trauma-related responses.

Conversely, health behaviors—such as consistent mask-wearing, hand hygiene, and timely medical consultation—are essential protective responses during pandemics. These behaviors not only reduce physical risk but may also serve as psychological buffers, fostering a sense of control and reducing anxiety. In the context of PTSD, positive health behaviors may mediate or moderate symptom severity by reinforcing structure and agency amid chaos.

Importantly, although both experiential avoidance and health behaviors are considered modifiable predictors of PTSD symptoms, their effects are not uniformly distributed across individuals. These effects may become conditional, varying in strength or direction depending on sociodemographic moderators such as age, gender, or professional role. For instance, avoidance tendencies may exert stronger effects on PTSD among younger HCWs with less coping capacity, while the protective influence of health behaviors may be more pronounced among older or more experienced personnel. Identifying these conditional pathways is crucial for developing targeted interventions that address trauma risk across diverse healthcare worker populations.

On the other hand, the Health Belief Model (HBM) offers a theoretical lens through which the relationship between avoidance tendencies, health behaviors, and psychological outcomes can be examined. Traditionally, the HBM has emphasized individuals’ perceptions of susceptibility, severity, benefits, and barriers to health actions [14–18]. However, Strecher, Champion, and Rosenstock [19] criticized the routine implementation of HBM as a simplistic four-variable model, urging researchers to explore more complex causal pathways. In line with this recommendation, HBM can be reconceptualized as a mediating mechanism, helping to explain how psychological avoidance and behavioral responses interact to influence trauma-related outcomes such as PTSD. Despite its conceptual relevance, there remains a notable gap in the empirical literature applying this model to trauma experiences among healthcare workers in high-risk African settings. In examining this within the Nigerian context, it is essential to consider how socio-demographic variables may further shape these trauma-related processes.

In Nigeria, socio-demographic variables such as age, professional designation, and years of experience carry significant weight, both culturally and institutionally. Culturally, age symbolizes wisdom and dignity, yet in formal systems such as the civil service or healthcare institutions, authority is more often determined by years of service and professional rank. In healthcare specifically, professional designation dictates hierarchy and decision-making power; medical doctors may overlook or minimize the contributions of laboratory scientists, pharmacists, or physiologists. These imbalances can lead to emotional strain and workplace invalidation, particularly for lower-status professionals. Years of practice also shape trauma exposure, resilience, and coping mechanisms. While variables like sex and education remain relevant, they may not reflect the embedded institutional dynamics that shape psychological vulnerability in this context. Despite growing interest in trauma-informed care, limited research has examined whether the indirect effects of trauma-related behaviors on PTSD—via mediators such as the Health Belief Model (HBM)—vary by these socio-demographic factors. Addressing this gap, the current study focuses on age, designation, and years of professional experience as key moderators of the trauma–PTSD pathway.

Also, Sociodemographic factors—such as age, gender, and professional role—may function as important moderators in the relationship between experiential avoidance, health behaviors, and PTSD outcomes. While these variables have often been treated as static predictors in previous studies, their potential to shape psychological vulnerability and resilience is frequently overlooked. This tendency has led to generalized conclusions about their influence, without adequately examining their modifiability or interaction with contextual stressors. For instance, younger healthcare workers or those in junior professional roles may exhibit different coping patterns or risk levels compared to their older or more senior counterparts. A more nuanced understanding of these moderators can contribute to the development of tailored interventions that reflect the specific needs and risk profiles of different subgroups within the healthcare workforce. Recognizing these variables as dynamic moderators, rather than fixed predictors, allows for a more contextually sensitive approach to understanding PTSD risk and resilience within high-stress healthcare environments.

Taken together, this study advances a conceptual model in which experiential avoidance and health-related behaviors exert both direct and indirect effects on PTSD symptoms among healthcare workers. The Health Belief Model (HBM) is proposed as a theoretical mediator, elucidating the psychological mechanisms linking avoidance tendencies to trauma-related outcomes. Sociodemographic characteristics—such as age, gender, and professional role—are further hypothesized to moderate these pathways, influencing differential susceptibility or resilience. By examining these interrelated constructs within the context of a high-risk African setting, this study aims to deepen our understanding of the psychological sequelae of public health crises and inform culturally and contextually relevant interventions for frontline healthcare workers.

While the COVID-19 pandemic triggered significant psychological distress among healthcare workers worldwide, little is known about the specific psychological processes driving trauma responses among frontline workers in resource-constrained and socially unstable settings such as Ekiti State, Nigeria. Despite mounting evidence of post-traumatic stress disorder (PTSD) among healthcare workers globally, existing research in Nigeria has largely overlooked key cognitive-emotional mechanisms, such as experiential avoidance, that may underlie PTSD symptoms in such environments.

The pandemic posed not only a biological threat but also a profound psychological challenge for frontline healthcare workers, particularly during its early stages marked by uncertainty, misinformation, and fear. In Ekiti State, where health systems are often under-resourced and workers operate under immense pressure, the emotional and cognitive burdens borne by healthcare providers merit urgent scholarly attention.

This study aims to investigate the psychological risk factors contributing to PTSD symptoms among healthcare workers during early-stage pandemics, with a particular focus on the role of the Health Belief Model (HBM) and experiential avoidance (EA). It also explores how demographic factors such as marital status, educational attainment, and types of healthcare settings moderate these relationships.

While numerous prior studies have employed additive models to examine PTSD risk factors, this research advances the field by applying a conditional process framework—specifically moderated mediation and conditional mediation models. This approach allows for a more nuanced understanding of how experiential avoidance and health behaviors influence PTSD through health beliefs, and how these relationships vary across sociodemographic groups. Such complexity is critical to capturing the realities faced by healthcare workers in under-resourced settings where structural and psychological factors interact to shape trauma responses. The insights gained have the potential to inform tailored interventions and policies addressing both individual and systemic determinants of mental health among frontline healthcare workers. This study uniquely integrates the Health Belief Model and experiential avoidance to deepen understanding of PTSD risk among frontline healthcare workers, providing insights to guide targeted mental health interventions in resource-limited settings.

To address these aims, the following objectives were formulated:

1. To examine the direct relationships among experiential avoidance, health behaviors, health beliefs, and PTSD symptoms among healthcare workers.
2. To investigate whether health beliefs mediate the relationships of experiential avoidance and health behaviors with PTSD symptoms.
3. To evaluate the joint influence of experiential avoidance and health behaviors on PTSD through health beliefs.
4. To determine which specific components of health beliefs mediate the relationship between demographic or psychological factors and PTSD symptoms.
5. To examine whether personal and professional characteristics (such as age, designation, and years of practice) moderate the indirect effects of health behaviors and experiential avoidance on PTSD symptoms via health beliefs.
6. To investigate whether socio-demographic characteristics (e.g., age, sex, education, years of practice, and designation) moderate the direct effects of health behaviors, experiential avoidance, and health beliefs on PTSD symptoms.

## Significance of the Study

### PTSD and HBM

The outbreak of a novel and untreatable infection, such as COVID-19, can be considered a traumatic experience due to its acute and chronic impact on individuals, particularly healthcare workers across hospitals globally. Post-Traumatic Stress Disorder (PTSD) is a mental health condition that may develop after exposure to life-threatening or highly distressing events. It is characterized by emotional and psychological reactions to trauma, primarily involving re-experiencing, avoidance, and hypervigilance [10, 20]. PTSD is classified as an affective disorder marked by anxiety and fear, which can contribute to depression, helplessness, memory impairment, and diminished work performance [10]. The COVID-19 pandemic, classified as a natural disaster, holds the potential to generate widespread psychological harm. Among the mental health consequences, PTSD is often regarded as the “signature” psychiatric outcome of trauma [13, 21]. It has been widely studied in the aftermath of various disasters and consistently emerges as the most frequently diagnosed psychiatric disorder in such contexts [22]. As described above, the COVID-19 burden is associated with PTSD symptoms among health care workers.

As a public health emergency, the suddenness, unpredictability, development uncertainty, and complexity of COVID-19 caused public panic disorder, affecting people’s everyday lives, work order, and social stability [23–24], led to changes in people’s lifestyles and increased psychological burden, and further aggravated the occurrence of PTSD symptoms. Studies have shown that the types of symptoms associated with the pandemic (Post-COVID Stress Disorder) matches the criteria defined for PTSD and comprises hyperarousal (increased reactivity to stimuli), avoidance (symptoms manifested in evading stimuli associated with the event), re-experiencing (**recurrent and distressing memories, flashbacks, nightmares** (which aggravates the sense of helplessness and depression) [25], as well as the dysphoric and anxious arousal associated with memories of the COVID-19 pandemic or related event [26–27]. In a study conducted by North et al [27] to provides an in-depth nosological consideration of the diagnosis of PTSD showed that the prevalence of PTSD ranges from 12% to 96% in hospitalized COVID-19 patients, 4% to 73% in healthcare workers and 3% to 67% of general populations.

North et al. [28] have criticized the use of the DSM-IV, DSM-5, DSM-5-TR, and ICD-11 in addressing the mental health consequences of the COVID-19 pandemic, arguing that these diagnostic frameworks are insufficient. They highlight conceptual issues within these diagnoses and call for empirical research to resolve diagnostic uncertainties. According to their perspective, understanding PTSD requires a clear distinction between three key concepts: the nature of the traumatic event, the individual’s exposure to it, and their emotional and psychological responses. The first two elements correspond to Criterion A in the DSM-5, while the third encompasses the symptom criteria (B–H). Ensuring a clear differentiation among these concepts is crucial to avoid confusion in trauma-related research and diagnosis.

While North et al. [28] argue for a clearer conceptualization of PTSD by distinguishing trauma, exposure, and emotional responses, the COVID-19 pandemic has accentuated the challenges of applying traditional diagnostic criteria to novel global crises. According to DSM-5, a traumatic event must involve direct exposure to actual or threatened death, serious injury, or sexual violence [29]. Although the pandemic is undeniably severe and potentially fatal, it is classified as a serious medical stressor rather than a traumatic event, and thus does not inherently qualify for a PTSD diagnosis under the DSM-5 criteria. However, the pandemic has given rise to a host of secondary stressors—economic hardship, social isolation, and loss of normalcy—that profoundly impact mental health. These nuances expose the limitations of DSM-5 in capturing the psychological impact of large-scale public health emergencies, particularly as they relate to the secondary trauma experienced by HCWs and the public.

Despite these definitional constraints, many studies have reported elevated PTSD symptoms during the pandemic, further fueling the debate about whether such symptoms arise from qualifying traumatic experiences. For example, a national survey in Ireland found that 17.7% of respondents met criteria for probable COVID-19-related PTSD (Karatzias et al. [30]. However, the study did not confirm whether participants had experienced events meeting DSM-5 Criterion A, nor did it assess for prior trauma exposure. This omission raises questions about whether the reported symptoms stem from trauma or from broader psychological distress. In contrast to DSM-5’s narrower criteria, the DSM-IV offers a broader and more inclusive definition of trauma, which recognizes emotional distress and vicarious trauma, thus allowing for the recognition of COVID-19-related stress as legitimate grounds for a PTSD diagnosis[10]. While ICD-11 provides a similarly inclusive definition, experts such as North et al. [28] and Van Overmeire [31] caution against broadening the trauma category too much, asserting that only individuals directly exposed to life-threatening COVID-19 events—such as ICU admissions or witnessing deaths—would meet the diagnostic threshold for PTSD. Thus, distinguishing between general distress and trauma-related psychopathology remains critical for maintaining diagnostic accuracy and avoiding the over-pathologization of normal stress responses.

Although more recent iterations of the DSM (DSM-IV and DSM-5) have introduced changes to the diagnostic criteria for PTSD, particularly Criterion A, these revisions have faced substantial critique. North et al. [27] argue that DSM-5 conflates the definition of trauma with its modes of exposure, thereby compromising diagnostic clarity. This entanglement is especially problematic in the context of the COVID-19 pandemic, where healthcare workers (HCWs) may experience trauma through direct and indirect exposures that do not always neatly fit DSM-5’s stricter criteria. In contrast, the DSM-IV provides a more distinct delineation between trauma and exposure, allowing for greater flexibility in identifying trauma-related symptoms in HCWs who experienced high levels of stress, vicarious trauma, and occupational hazard during the pandemic. The DSM-IV framework also more comfortably accommodates experiences of secondary trauma, which are prevalent among HCWs exposed to traumatic events through others’ suffering or indirect involvement, making it a more suitable choice for understanding the pandemic’s impact on mental health.

Therefore, this study adopts the DSM-IV framework, in alignment with North’s critique, to ensure both conceptual clarity and contextual relevance for examining PTSD symptoms among healthcare professionals during the COVID-19 pandemic. Several factors have contributed to the development of PTSD symptoms among healthcare workers (HCWs) during this time, with studies revealing that trauma experienced by HCWs often extends beyond direct exposure to life-threatening events. For example, in a study conducted among Chinese HCWs, Chen et al. [32] found that their primary concerns included the fear of infecting family members, difficulty managing patients’ panic, and shortages of protective equipment. These factors, alongside the perception of COVID-19 as an unpredictable and potentially fatal disease, have contributed to elevated PTSD symptoms, particularly when combined with other stressors such as isolation and stigma, which are key to understanding PTSD in healthcare settings. These findings were supported by Lai et al. [33], who surveyed 1,257 healthcare professionals working in COVID-19 wards across 34 hospitals in China. Similarly, an online survey conducted in Greece involving 270 HCWs [34] revealed high PTSD scores, which were associated with increased anxiety about the progression of the virus. Many HCWs perceived COVID-19 as an unpredictable and terminal disease, a perception influenced by limited experience with pandemics and inadequate resources. This perceived lack of control was found to be a strong predictor of post-traumatic stress symptoms (PTSS), leading to increased burnout and a diminished sense of professional competence [35]. Another contributing factor to PTSD development is the sense of isolation experienced by HCWs who were quarantined after exposure to the virus or who had recovered from infection themselves [36–37]. These individuals often reported feelings of stigma and self-isolation, driven by fears of transmitting the virus to loved ones. In addition a study from Slovakia during the COVID-19 pandemic indicated that re-experiencing in the form of rumination in connection to anticipated loss of income, insufficient opportunities to talk with others, and a subjective sense of isolation, low tolerance of uncertainty as well as catastrophizing, analyzed as psychological variables, decreased the frequency of positive experiences and increased the frequency of negative emotions and severity of depression [37]. These studies point that anxiety of the pandemic and experiencing the events as frontline healthcare workers as well as stressors associated with one family members and the environments and the memories of seeing patient dying helplessly exacerbated PTSD. However, no study has associated PTSD with health belief model (HBM).

The Health Belief Model (HBM), a psychological theory that originated in the United States, is one of the most widely used frameworks for understanding health-related behaviors. As the name suggests, the HBM posits that individuals are more likely to take preventive health actions if they perceive themselves as susceptible to a condition (perceived susceptibility), believe the condition has serious consequences (perceived severity), and view the recommended action as beneficial with minimal barriers (perceived benefits and perceived barriers) [18]. Initially developed to address the underutilization of preventive health services such as tuberculosis screening, the model emphasizes the importance of individuals’ cognitive assessments in guiding behavior [38, 16, 39]. For the purpose of this study, we focus on the model’s original components—perceived susceptibility, perceived severity, and perceived barriers.

This theoretical framework complements prior research showing that while demographic characteristics such as age, gender, education, and socioeconomic status significantly influence preventive health behaviors, these traits are largely unchangeable. However, as Conner and Norman [40] note, psychological factors like risk perception and health beliefs—which are central to the HBM—can be modified through targeted health education. By integrating insights from the HBM, health researchers and policymakers can develop strategies that transcend fixed demographic constraints and instead focus on shifting beliefs and perceptions to foster behavioral change at the population level.

Theoretical frameworks such as the Health Belief Model and trauma-related stress models have been used to understand how individuals respond to health threats under conditions of uncertainty and limited resources. During the early phase of the COVID-19 pandemic in Nigeria—particularly in Ekiti State—these frameworks became especially relevant. Public health messaging was often met with skepticism, especially among communities facing socioeconomic hardship and longstanding mistrust in government institutions [24–25]. These factors not only shaped individual responses to preventive behaviors but also influenced how healthcare workers navigated their roles on the frontlines.

Healthcare workers (HCWs) during this period were exposed to unusually high workloads, shortages of protective equipment, and significant emotional strain—all of which are known risk factors for psychological distress, including PTSD [14,16,18]. Psychological variables such as **experiential avoidance** may have influenced how HCWs coped with stress, while structural factors like **designation**, **years of practice**, and **educational attainment** may have moderated these effects. Given the combined influence of cognitive, emotional, and contextual factors, interaction effects among these variables warrant theoretical and empirical attention. This study thus draws on multiple frameworks to explore how both **individual psychological processes** and **sociodemographic characteristics** jointly influence health behavior and mental health outcomes in high-risk professional settings.

As observed by Champion & Skinner [41], HBM constructs are conceptualized in the literature as channels of influence of which we believed in the earlier stage of the pandemic, it could influence PTSD. Perceived susceptibility beliefs are deeply rooted in the personality of the individual, and they’re usually internalized during by healthcare workers and create emotional stability. Epidemiological <u>seroprevalence</u> studies consistently demonstrated that healthcare workers are at a higher risk of SARS-CoV-2 transmission compared to the general public [42–43]. In addition, lack of clarity regarding SARS-CoV-2 transmission patterns [44]and the difficulty of implementing guidelines issued by international governing bodies that are continuously changing [445–46]. COVID-19 is mainly transmitted when people breathe in air contaminated by droplets/aerosols and small airborne particles containing the virus. Infected people exhale those particles as they breathe, talk, cough, sneeze, or sing [47–51]. Transmission is more likely the closer people are. However, infection can occur over longer distances, particularly indoor[47, 51].

Other studies in UK and US comprising 99,795 healthcare workers self-reported COVID-19 data through a mobile application regarding COVID-19 risk factors and PPE usage. They found that even among healthcare workers who had access to adequate PPE, there was an increased susceptibility to COVID-19 infection [52]. They also found that adequate availability of PPE did not completely reduce the risk of infection in healthcare workers caring for COVID-19 patients and that reusing PPE was positively associated with an increased risk of infection for healthcare workers caring for COVID-19 patients [52]. With this high level of susceptibility, predict that HCWs workers in Ekiti state who relaxed on mode of transmission from scientists from the developed nation could not only lead to high level of susceptibility but also severity which could as influenced PTSD. Research have reported that individuals with high perceived susceptibility are likely to take relevant actions to reduce level of risk, while those with low perceived susceptibility are not likely to engage in health promotion behavior to mitigate the level of perceived danger [53]. At the same time Perceived severity known as perceived consequences or seriousness as well as perceived impediment (barriers) could lead to emotional distress and lead to psychological infliction such as PTSD. Yarseah et al [6] demonstrated that perceived severity and barriers predicted PTSD although perceived susceptibility could not.

While a veteran study emphasizes how health beliefs influence treatment behavior such as medication use, therapy participation in individuals already diagnosed with PTSD [54], the present study adopts a different lens. Specifically, it investigates how health beliefs function as emotionally charged cognitive factors—such as perceived severity of the virus, susceptibility to infection, and perceived barriers to safety or care—which may exacerbate the development or intensity of PTSD symptoms in healthcare workers (HCWs) during the COVID-19 pandemic. In this way, health beliefs are conceptualized not as predictors of treatment engagement, but as psychological stressors in their own right that potentially contribute to trauma-related psychopathology. This aligns with the Health Belief Model (HBM) by emphasizing how emotionally laden appraisals of risk and threat can shape mental health outcomes. Thus, whereas prior research connects health beliefs to post-diagnosis behavior, this study extends the model by exploring how such beliefs may influence the onset or severity of PTSD itself, particularly among HCWs functioning under extreme public health pressures.

### Experiential avoidance (EA) and Posttraumatic stress disorder (PTSD)

Experiential avoidance, (EA) is a problem that happens when an individual refused to worry or think about the issues and take all necessary steps to avoid contact with any resemblance of the phenomenal. EA, as an inhibitory coping strategy for emotion regulation, is associated with a range of psychiatric disorders, including anxiety and depression [12]. In the case of the coronavirus pandemic, a nurse could avoids thinking or talking about the number of patients who died during her shift, or a nurse experiences chest tightness and palpitations due to anxiety but avoids seeking medical help for fear of being diagnosed with COVID or appearing weak. Experiential avoidance includes any behavior aimed eluding unpleasant internal experiences or the external circumstances that trigger them [13].These avoided experiences may encompass thoughts, emotions, physical sensations, or other distressing internal states. Experiential avoidance does not specify a particular form or manifestation of behavior but rather represents a broad category of behaviors united by their common goal of escaping unwanted internal experiences [21].

There are many studies that have confirmed relationship between EA and PTSD [12–13, 21, 6]. In particular, the study confirms a significant positive association between experiential avoidance (EA) and PTSD individuals who engage more in avoiding painful thoughts, memories, or emotions are more likely to experience PTSD symptoms [55]. In some studies, EA is seeing as a deliberate self-harm [56] and by extension, EA is understood to be a transdiagnostic psychological construct relevant to several mental health problems, such as depression, substance use, anxiety-related disorders, and PTSD [57–58]. This supports previous literature and theoretical frameworks suggesting that EA is not just a coping style but a potential risk factor for the development and maintenance of PTSD [11–13]. Avoidance may offer temporary relief, but over time, it interferes with trauma processing and contributes to symptom persistence.

While experiential avoidance (EA) is often conceptualized as a maladaptive coping strategy, it can also serve adaptive functions in high-risk or life-threatening situations, such as during the COVID-19 pandemic. For instance, a healthcare worker experiencing intense fear or anxiety about contracting the virus may temporarily redirect their attention toward immediate tasks— such as monitoring vital signs or administering treatments—to remain functional. In such contexts, avoiding distressing thoughts about personal vulnerability may help prevent emotional overwhelm and allow continued performance of critical duties. This short-term form of EA may therefore function as an **autonomous, emotion-regulating strategy**, enabling healthcare workers to protect themselves psychologically in the moment and avoid catastrophic breakdowns. Although this self-protective use of EA may offer temporary relief, its prolonged use can impede trauma processing and contribute to persistent PTSD symptoms over time [59]. While this adaptive dimension of EA has received limited attention in previous research, it is essential to consider its dual role—**as both a potentially functional short-term coping strategy and a risk factor for long-term psychological distress**. Existing studies have not adequately explored the dynamic relationship between EA and PTSD among frontline healthcare workers during the COVID-19 pandemic, particularly in resource-constrained contexts like Nigeria, revealing an important gap that this study aims to address [12–13,58,60].

### Health Belief Model and Experiential avoidance

The Health Belief Model (HBM) emphasizes individuals’ mental models regarding health and health behavior, focusing on two main components: threat perception and behavioral assessment. Threat perception involves beliefs about how susceptible one is to a health issue and how severe the consequences of that issue might be. Behavioral assessment includes beliefs about the benefits or effectiveness of taking a recommended health action, as well as the potential barriers or costs associated with it [61] For examples, during the COVID-19 pandemic, individuals who perceived themselves as highly susceptible to infection—such as frontline healthcare workers or the elderly—were more likely to adopt preventive behaviors. Those who believed the virus could lead to severe health consequences, including hospitalization or death, also showed greater compliance with safety measures like vaccination and mask-wearing. However, perceived barriers such as vaccine side effects, misinformation, and discomfort with masks often discouraged people from engaging in recommended health actions. However the effectiveness of health belief model has often been question. Carpenter[16]noticed that going through a review of the effectiveness of the Health Belief Model to predict and explain behavior found that perceived severity was a weak predictor of health behavior, perceived susceptibility was not a predictor in most studies, and perceived barriers and perceived benefits were consistently the strongest predictors of health behavior [16]. But we do not know the relationship between HBM and experiential avoidance.

Experiential avoidance fundamentally embodies repression and situational escape or suppression which is an active effort to control or eradicate immediate experiences of negative private events, such as unwanted thoughts, feelings, memories, or physical sensations. Situational escape/avoidance involves attempting to modify contextual factors that are likely associated with the occurrence of undesirable internal experiences[13] Therefore, experiential avoidance includes any behavior aimed eluding unpleasant internal experiences or the external circumstances that trigger them[59] These avoided experiences may encompass thoughts, emotions, physical sensations, or other distressing internal states. Experiential avoidance is known for being adaptive in a short time providing a way to manage overwhelming emotions and stay functional during a crisis like the COVID-19 pandemic. However, excessive cognitive, emotional, and behavioral avoidance necessitates significant time and energy for management and control, leading to disordered processes. Moreover, excessive avoidance impedes progress toward valuable goals and limits available experiences [12]

### Mediating roles of HBM on EA and PTSD

In the early stages of the COVID-19 pandemic, many African healthcare workers (HCWs) experienced significant emotional distress due to uncertainty, lack of information, and inadequate preparedness. Media reports highlighting high mortality rates among HCWs in Europe and Asia likely intensified perceptions of susceptibility and severity, while perceived barriers such as limited access to personal protective equipment (PPE) and delayed testing results heightened feelings of helplessness. Studies suggest that, under these conditions, the Health Belief Model (HBM), traditionally used to predict health behaviors, may have contributed more to psychological strain than to the intended protective behaviors [6]. This is partly due to its limited consideration of emotional and contextual factors, as well as its assumption that individuals make purely rational decisions even in high-stress or trauma-laden situations[62–63]. Furthermore, experiential avoidance (EA), a coping mechanism where individuals evade distressing emotions and thoughts, likely interacted with these health beliefs, exacerbating the risk of post-traumatic stress disorder (PTSD). Therefore, the HBM may mediate the relationship between EA and PTSD by shaping how HCWs cognitively and emotionally respond to the crisis but to this date, to our knowledge, there is no research that has investigated the the mediating roles of HBM on PTSD.

Research from past pandemics, including H1N1 and COVID-19, has shown that health-protective behaviors are influenced not only by external public health messaging but also by internal belief systems grounded in HBM constructs [18].Indeed healthcare workers (HCWs) in Africa experienced exceptionally high levels of perceived susceptibility and severity, driven by the rapid spread of the virus and limited access to critical medical resources. This gap between health beliefs and behavior may be better understood through the lens of the Health Belief Model (HBM), which emphasizes the role of perceived barriers—such as inadequate supply chains, limited training, or stigma around psychological support—in shaping health-related decision-making. In this context, HBM constructs may mediate the relationship between experiential avoidance and PTSD symptoms, highlighting how internal belief systems and environmental limitations interact during public health crises

This disconnect may be partly explained by experiential avoidance, the tendency to evade or suppress distressing internal experiences, including anxiety, fear, shame, or trauma-related thoughts and emotions [12–13]. EA can distort the functionality of the HBM in several ways. It may reduce the emotional salience of perceived susceptibility and severity (“I know I’m at risk, but thinking about it overwhelms me, so I avoid it”), inflate perceived barriers (e.g., “Talking about what I’ve seen will make me break down—I can’t handle that”), and diminish the perceived benefits of action (e.g., “Using PPE constantly reminds me of how bad things are—it’s easier not to think about it”).

Research suggests that experiential avoidance (EA) may disrupt the link between health beliefs and protective behaviors. For instance, healthcare workers may acknowledge their vulnerability and recognize the benefits of preventive strategies, yet emotionally disengage from these actions due to the psychological distress they elicit. This avoidance has been associated with reduced coping behaviors and prolonged exposure to traumatic stressors, ultimately increasing vulnerability to PTSD symptoms [57,59].

Although meta-analytic reviews [64–65] have critiqued the Health Belief Model (HBM) for its modest predictive utility, more recent evaluations [16] suggest that certain constructs, particularly perceived barriers and benefits, consistently influence behavior. Rather than using HBM strictly as a predictive model, this study conceptualizes it as a **mediating psychological framework—**one through which **experiential avoidance may influence the development of PTSD symptoms**. In the emotionally and resource-constrained context of the COVID-19 pandemic, this framework allows for a more nuanced understanding of how healthcare workers’ internal processes and belief systems interact to produce trauma-related outcomes.

The proposed mediation model addresses a critical gap in existing literature by emphasizing how emotional regulation difficulties in the form of EA interacts with cognitive health beliefs to shape trauma-related outcomes. By integrating experiential avoidance with HBM constructs, this study offers a more comprehensive understanding of why many healthcare workers, despite high awareness of risks, fail to engage in self-protective behaviors. It also sheds light on how this dissonance between belief and behavior may contribute to long-term psychological consequences during high-risk public health emergencies.

### Moderation roles of some Sociodemographic Variables on the mediating effects of Health belief model

The Health Belief Model (HBM) is a foundational framework in health psychology explaining how individuals’ beliefs about health threats influence their behaviors [66]. It assumes rational decision-making, where individuals act to avoid negative health outcomes if they believe the action is effective and feasible. However, this assumption may not hold in all contexts, particularly during acute crises like the COVID-19 pandemic, when individuals may disengage from health information or avoid protective behaviors despite risks ([61, 63].

Traditionally, HBM research employs an additive approach, often overlooking the complex interplay between psychological processes and personal background variables such as age, gender, or profession. Strecher, Champion, and Rosenstock [19] highlighted the need to explore how background characteristics interact with belief structures to influence behavior. Addressing this gap, the current study applies a parallel mediation model that views health beliefs as distinct pathways—such as perceived susceptibility, severity, and barriers—through which psychological factors like experiential avoidance affect PTSD symptoms.

Recent research extends HBM beyond predicting behavior, considering it as a mediating mechanism linking psychological and behavioral factors to mental health outcomes (e.g., PTSD) [15–16]. Moreover, moderation, where third variables influence the strength or direction of relationships is increasingly recognized in health psychology, with sociodemographic factors playing key roles [18,60,13]. This study integrates these insights by employing a moderated mediation framework, capturing the nuanced, conditional effects of health beliefs and individual differences on trauma responses among healthcare workers.

This study will investigate whether sociodemographic variables—such as age, sex, education, professional designation, and years of practice—moderate the indirect relationship between experiential avoidance and PTSD symptoms, with health beliefs acting as a mediator. Through a conditional process approach grounded in the Health Belief Model (HBM), we aim to determine whether these mediating mechanisms vary across different demographic groups.

In healthcare, workers operate in cognitively demanding roles and serve as both providers and recipients of mental health care. Their interpretation of health information and engagement with services are influenced by personal and professional attributes. Notably, women constitute up to 70% of healthcare workers globally, especially in nursing ([67], yet gender’s moderating role in PTSD among healthcare workers remains underexplored [68–70], suggesting a critical research gap [70].

While prior studies have included sociodemographic variables as predictors of PTSD and related outcomes [70–75], these are often treated additively, overlooking their interactive effects with psychological and belief systems. Such simplification limits understanding of trauma responses in high-stress healthcare environments where individual and contextual factors modulate psychological mechanisms.

Modeling sociodemographic factors as moderators within a conditional process framework reveals how experiential avoidance’s impact on PTSD via health beliefs differs by subgroup, clarifying when and for whom these processes are most relevant. For example, studies show that higher educational attainment and health-related knowledge correlate with greater engagement in protective health behaviors during pandemics [76–78]. In a study in China, Kim and Kim [77] found that self-efficacy, female gender, and perceived severity were among the strongest predictors of preventive actions, while resource variables such as education level and social support moderated the effects of health beliefs on behavior. These findings emphasize the importance of integrating both belief-based and structural resource factors to fully understand individual-level compliance during health crises. While these studies have illuminated the role of health beliefs and resource factors in shaping preventive behavior during pandemics, they often overlook the emotional mechanisms and mental health consequences such beliefs may entail particularly among healthcare workers. By incorporating experiential avoidance and employing a conditional process model, the present study addresses these gaps, offering a more nuanced understanding of how cognitive and emotional factors interact to influence PTSD symptoms in a high-risk, underrepresented population.

Female healthcare workers and nurses experience disproportionately higher psychological distress [79–81]. COVID-19 research identifies gender, age, and marital status as key risk factors, with females reporting greater burden, while age and marital status effects vary [82–85]. Professional characteristics like years of practice and job role also predict PTSD symptoms [72–74, 86], with evidence of demographic and occupational differences in experiential avoidance and health beliefs during COVID-19 [6]. Previous research suggests that unmarried healthcare workers may show higher perceived severity and experiential avoidance, underscoring marital status as a potential moderator of psychological outcomes [83]. These findings highlight the importance of context and background in shaping trauma-related psychological outcomes.

## Methodology

### Ethical Statement

Ethical approval for this study was granted by the **Ekiti State University Ethical Review and Research Committee**, coordinated through the **Office of Research, Development, and Innovation (ORDI)**. The research protocol, which outlined the study’s objectives, methodology, and consent procedures, was reviewed and approved in accordance with the ethical standards set forth in the **Declaration of Helsinki**. Due to pandemic-related movement restrictions, the University Ethics Committee authorized the research to be conducted under the close oversight of the **Ekiti State Ministry of Health and Human Services**. The Ministry ensured adherence to the ethical principles of **respect for persons**, **justice**, and **beneficence** throughout the study. Regular updates were submitted to the Ministry, and no deviations from the approved protocol occurred. Upon completion of the study, a full report was submitted to the **Ekiti State University Ethical Review and Research Committee**, through the **Office of Research, Development, and Innovation (ORDI)**, for final review and issuance of the ethical clearance statement.

### Research Design

This research adopted a quantitative research design to build on the findings from the previous study and to refine our understanding of PTSD risk among healthcare workers during early-stage pandemics. The present study is distinct from the first by emphasizing the role of the Health Belief Model (HBM) and experiential avoidance (EA) as key constructs influencing PTSD risk. Additionally, demographic moderators such as age, gender, education level, and marital status are integrated to explore how these factors shape psychological responses to health threats.

While the first study examined healthcare workers’ health beliefs and coping strategies in an exploratory manner, this study applies a more focused analytical approach, using moderation analyses to investigate how demographic factors influence the HBM-EA-PTSD pathway. This allows for a deeper understanding of how different individual characteristics modify psychological outcomes related to trauma exposure and health beliefs.

Given the ongoing impact of the COVID-19 pandemic and the unique stressors faced by healthcare workers, the study aims to provide a more nuanced understanding of how experiential avoidance and health beliefs interact with demographic factors to affect PTSD risk. The same dataset, previously used to examine PTSD development, is re-analyzed through this new lens, contributing to the existing literature by expanding the scope of variables considered.

To ensure safety during data collection amidst the ongoing pandemic, we followed stringent health and safety protocols, including the use of personal protective equipment (PPE) and maintaining social distancing. Data collection was again carried out via surveys, but with a more targeted focus on the variables under investigation in this study.

### Setting and Participants

The study was conducted in Ekiti State, Southwest Nigeria, a region that continues to experience significant public health challenges, including the COVID-19 pandemic. Ekiti State was among the most affected areas in West Africa during the early stages of the pandemic, which influenced healthcare workers’ mental health. The research focused on urban areas within the state, particularly the Ado and Ido-Osi Local Government Areas (LGAs), which are home to the state’s major healthcare facilities. These urban settings were particularly critical as they housed the only three tertiary healthcare facilities in Ekiti State, which played a central role in treating COVID-19 patients during the pandemic’s early phase.

Ekiti State has a population of approximately 5.4 million, with a high concentration of healthcare facilities, including primary, secondary, and tertiary hospitals. For the purposes of this study, we focused on the healthcare workers in the two LGAs due to their significant healthcare infrastructure. These areas collectively have 104 hospitals, including 70 primary healthcare facilities, 31 secondary healthcare facilities, and the three tertiary healthcare facilities.

This study specifically engaged healthcare workers from these facilities, aiming to explore the role of demographic moderators (such as age range, designation, years of practice, sex, and education attainment) on the pathway from Health Belief Model (HBM) constructs to Experiential Avoidance (EA) and PTSD risk. The demographic data gathered in the first study is being re-analyzed to explore the moderating effects of these variables on the psychological outcomes related to the pandemic.

### Participants

The participants in this study were healthcare workers from the urban areas of Ado and Ido-Osi, including medical doctors, nurses, and other frontline professionals who were directly affected by the COVID-19 pandemic. The sample included a diverse range of individuals across various demographic categories. These demographic variables were examined as moderating factors in the relationship between experiential avoidance, health behaviors, health beliefs, and PTSD symptoms arising from pandemic-related exposure.

### Sample Size Determination

A post hoc power analysis was conducted using G*Power (version 3.1.9.7) to determine the adequacy of the sample size. With 8 predictors, an alpha level of 0.05, and a medium effect size (f² = 0.15) based on Cohen’s guidelines Cohen [87], a minimum of 109 participants was required to achieve a power of 0.80. Given the final sample size of 475, the study was adequately powered to detect even small effects (f² ≥ 0.02), enhancing the reliability of the structural path estimates.

### Sampling and Recruitment

A total of 475 healthcare workers in Ekiti State were recruited using snowball sampling, a strategy chosen to facilitate access to frontline workers under the highly restricted and sensitive conditions of the pandemic. Following appropriate consultation with the Ekiti State Medical Board, initial participants were identified through professional healthcare networks and were encouraged to refer eligible colleagues, promoting trust and increasing participation rates.

All participants provided informed consent, and ethical procedures were strictly followed to ensure confidentiality and data integrity. Data were collected between May 5th and June 18th, 2020, during a critical period of heightened exposure and uncertainty for healthcare workers.

As shown in Table 1, the study sample consisted of 475 healthcare workers, most of whom were aged between 25 and 50 years, with the largest age group being 25–30 years (25.5%). A majority were female (64.0%) and married (76.4%), suggesting a relatively young, predominantly female, and married workforce. Regarding professional designation, the highest proportion of respondents were nurses (37.3%), followed by doctors (22.7%) and other health professionals such as laboratory scientists, pharmacists, and CHEWs. The participants were fairly evenly distributed between primary healthcare centers (41.9%) and tertiary hospitals (40.4%), with fewer working in secondary healthcare settings (17.7%). In terms of ownership, most respondents were employed in public healthcare facilities (81.9%), reflecting the dominant role of government institutions in healthcare provision.

**Table 1:** Socio-Demographic Characteristics of Respondents (N = 475)

| Variable | Category | Frequency | Percent (%) | Valid Percent (%) | Cumulative Percent (%) |
| --- | --- | --- | --- | --- | --- |
| Age Range | 25–30 | 121 | 25.5 | 25.5 | 25.5 |
|  | 31–35 | 101 | 21.3 | 21.3 | 46.7 |
|  | 36–40 | 77 | 16.2 | 16.2 | 62.9 |
|  | 41–45 | 56 | 11.8 | 11.8 | 74.7 |
|  | 46–50 | 98 | 20.6 | 20.6 | 95.4 |
|  | 61–65 | 12 | 2.5 | 2.5 | 97.9 |
|  | 71–75 | 6 | 1.3 | 1.3 | 99.2 |
|  | 76–80 | 4 | 0.8 | 0.8 | 100.0 |
| **Sex** | Male | 171 | 36.0 | 36.0 | 36.0 |
|  | Female | 304 | 64.0 | 64.0 | 100.0 |
| **Marital Status** | Married | 363 | 76.4 | 76.4 | 76.4 |
|  | Single | 98 | 20.6 | 20.6 | 97.1 |
|  | Divorced | 2 | 0.4 | 0.4 | 97.5 |
|  | Widow | 12 | 2.5 | 2.5 | 100.0 |
| **Designation** | Doctor | 108 | 22.7 | 22.7 | 22.7 |
|  | Nurse | 177 | 37.3 | 37.3 | 60.0 |
|  | EHO | 14 | 2.9 | 2.9 | 62.9 |
|  | CHEW | 38 | 8.0 | 8.0 | 70.9 |
|  | Epidemiologist | 12 | 2.5 | 2.5 | 73.4 |
|  | Laboratory Scientist | 41 | 8.6 | 8.6 | 82.0 |
|  | Pharmacist | 18 | 3.8 | 3.8 | 85.8 |
|  | Cleaner | 5 | 1.1 | 1.1 | 86.9 |
|  | Psychologist | 12 | 2.5 | 2.5 | 89.4 |
|  | X-ray | 10 | 2.1 | 2.1 | 91.5 |
|  | Patient Transporter | 40 | 8.4 | 8.4 | 99.9 |
| **Types of Healthcare Settings** | Primary Health Center | 199 | 41.9 | 41.9 | 41.9 |
|  | Secondary Health | 84 | 17.7 | 17.7 | 59.6 |
|  | Tertiary Hospital | 192 | 40.4 | 40.4 | 100.0 |
| **Ownership** | Public | 389 | 81.9 | 81.9 | 81.9 |
|  | Private | 86 | 18.1 | 18.1 | 100.0 |
| **Department Unit** | Maternity | 172 | 36.2 | 36.2 | 36.2 |
|  | A&E | 38 | 8.0 | 8.0 | 44.2 |
|  | GOPD | 62 | 13.1 | 13.1 | 57.3 |
|  | Infectious Disease Isolation Center | 28 | 5.9 | 5.9 | 63.2 |
|  | Others | 175 | 36.8 | 36.8 | 100.0 |

Departmental distribution showed that the largest groups worked in maternity units (36.2%) and unspecified “other” departments (36.8%), followed by general outpatient departments (13.1%), accident and emergency (8.0%), and infectious disease isolation centers (5.9%). These figures highlight the prominence of maternal and general health services in the facilities surveyed.

#### Methods: Rationale for Moderator Selection

This study tested moderated mediation models to examine whether the indirect effects of health behavior and experiential avoidance (EA) on PTSD symptoms via the Health Belief Model (HBM) were contingent on specific socio-demographic factors. Age, designation, and years of professional experience were selected as moderators based on theoretical and empirical considerations. These variables were chosen because they are likely to influence how individuals perceive health threats, regulate avoidance behavior, and internalize trauma-related beliefs. Other variables, such as sex and education, were not included as moderators in this model because their influence was not expected to operate through the specific psychological pathways being tested.

### Assumption Testing

Before conducting Structural Equation Modeling (SEM), assumptions for multivariate analysis were thoroughly assessed. The data were approximately normally distributed, with skewness and kurtosis values within ±2—an acceptable threshold supported by Kline and Tabachnick & Fidell [88–89]. Multicollinearity was not a concern, as all VIF values were below 5, aligning with guidelines recommended by Hair et al. and O’Brien [90–91]. No extreme outliers were detected, and missing data were minimal and appropriately handled.

### Justification of Statistical Tests

The use of Partial Least Squares Structural Equation Modeling (PLS-SEM) version 4.1 was justified given its ability to manage complex models and its robustness against non-normality, as emphasized by Hair et al. [92]. Normality was not a strict assumption, since PLS-SEM relies on non-parametric bootstrapping for estimating significance levels [93–94]. A bootstrapping procedure with 10,000 resamples was implemented to ensure reliable and stable estimates, consistent with methodological best practices in PLS-SEM [95].

## Result

The descriptive statistics for the study variables revealed varying levels of responses across participants. The mean score for perceived susceptibility was 33.12 (SD = 6.058), indicating that participants generally reported a moderate level of susceptibility, with some variability. For perceived severity, the average score was 23.90 (SD = 5.222), suggesting that participants perceived the condition to be moderately severe, with responses varying slightly. The perceived barriers were higher, with a mean of 44.91 (SD = 7.334), reflecting a significant variation in the barriers participants experienced. Psychological flexibility had a mean score of 14.43 (SD = 8.040), indicating moderate levels of flexibility, with considerable variation in responses. Participants’ PTSD scores averaged 30.34 (SD = 12.054), showing moderate symptoms with substantial variation. For re-experiencing, the mean was 9.39 (SD = 4.362), indicating moderate levels of re-experiencing symptoms. The average score for avoidance was 11.16 (SD = 4.901), pointing to moderate avoidance behaviors with some variability. Hyper-vigilance showed a mean of 9.79 (SD = 4.209), reflecting moderate levels of hyper-vigilance symptoms, with a degree of variation in responses. Finally, experiential avoidance had the same mean of (M = 14.43, SD = 8.040), with variability suggesting differing levels of avoidance among participants.

As shown in Table 3, there are significant correlations between the key variables in the study. Notably, PTSD symptoms are strongly correlated with re-experiencing (r = 0.891), avoidance (r = 0.908), and hypervigilance (r = 0.883), suggesting that these trauma-related symptoms are closely tied to the severity of PTSD. Psychological flexibility also shows moderate positive correlations with these PTSD-related symptoms, indicating that individuals with greater flexibility may experience less severe trauma symptoms. Additionally, perceived barriers to health and experiential avoidance were positively correlated with PTSD, re-experiencing, and avoidance, while showing a negative relationship with psychological flexibility.

### Measurement Model Evaluation

The first step in SmartPLS analysis involves assessing the measurement model. After conducting data screening and cleansing using Mahalanobis distance (p < .02) to check for outliers, attention was focused on the research model. The measurement model evaluation involved assessing composite reliability (Rho A), composite reliability (CR), Cronbach’s Alpha, convergent validity, and discriminant validity [96].

In examining the measurement model, the first priority was to assess **indicator reliability**. Indicator reliability refers to the proportion of indicator variance explained by the underlying construct [97]. This is typically measured through **outer loadings** [98], with loadings greater than 0.70 considered desirable [99–100]. However, it is common in social science research for some outer loadings to fall below 0.70. In such cases, indicators with loadings between 0.40 and 0.70 should only be considered for removal if doing so improves Composite Reliability (CR) or Average Variance Extracted (AVE) [99]. Accordingly, as shown in Table 2, two items (D38 and Q10) were removed because their outer loadings were below 0.50, following the recommendation of Green and Straub [90].

**Table 2:**
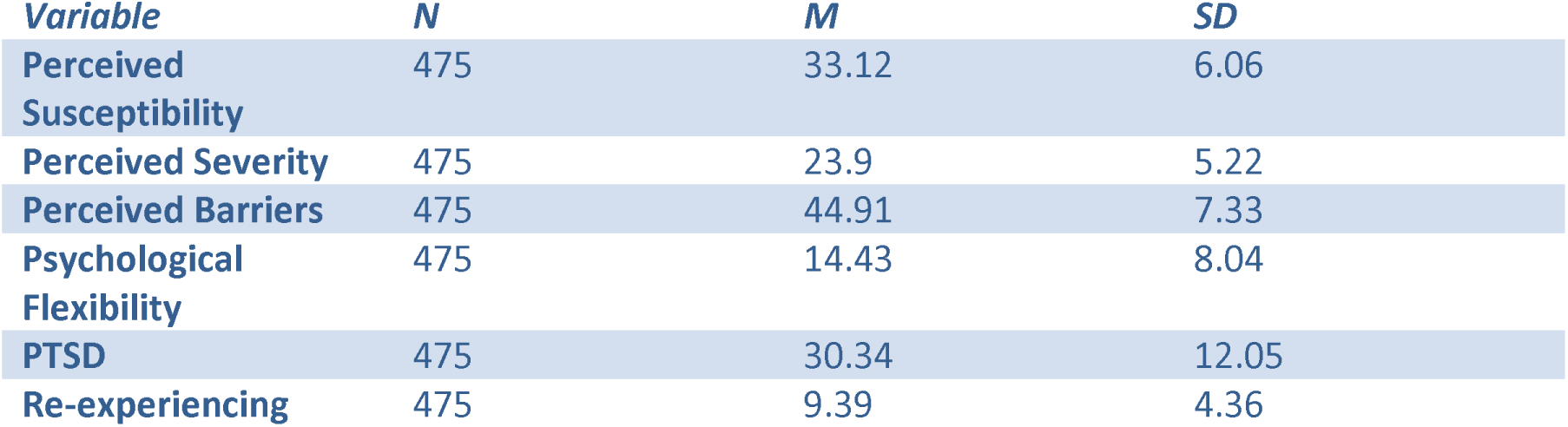
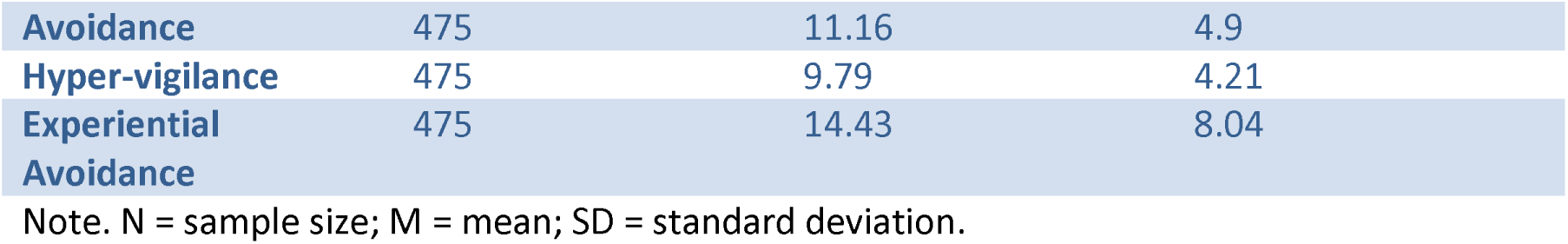
Descriptive Statistics for Key Variables.

**Table 3:**
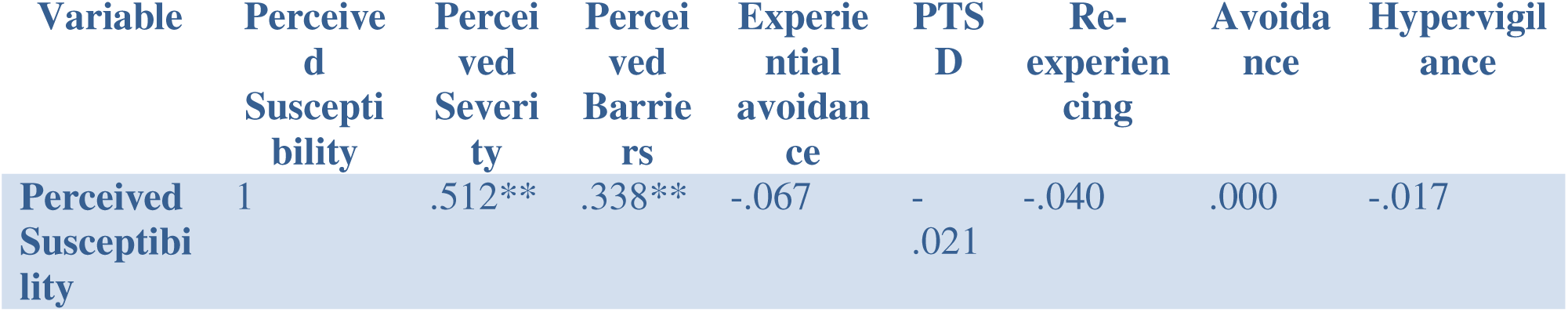
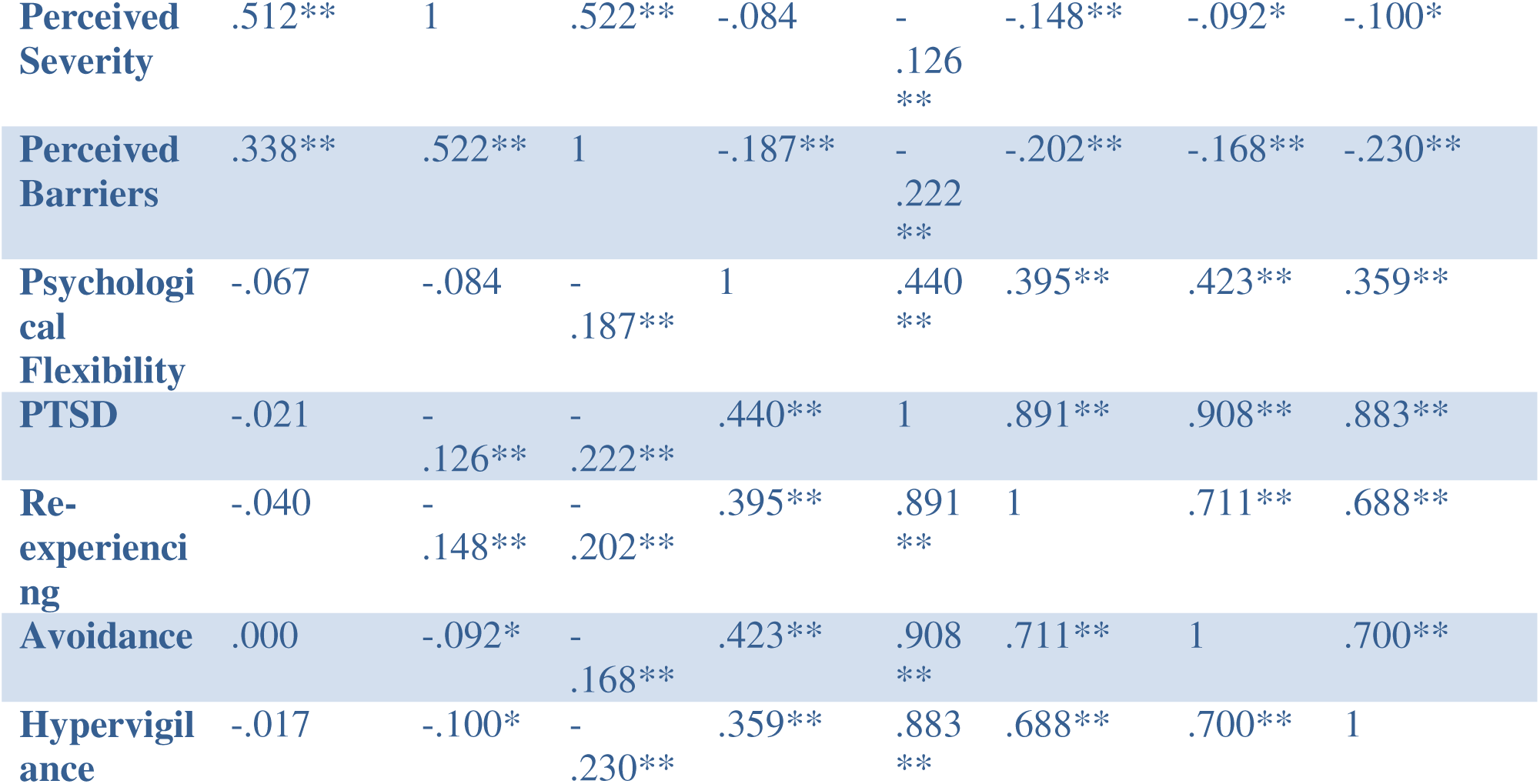
Socio-demography Variables Correlation Matrix.

**Construct reliability** was assessed using **Cronbach’s Alpha**, **Rho_A**, and **Composite Reliability (CR)**. Most constructs exceeded the recommended threshold of 0.70. However, the **Health Behavior** construct had a lower Cronbach’s Alpha. Despite this, both **Composite Reliability (CR)** and **Average Variance Extracted (AVE)** were within acceptable ranges, suggesting adequate reliability. It is important to note that **Cronbach’s Alpha** assumes **tau-equivalence** (equal factor loadings), which is often an unrealistic assumption in applied research [101–102]. **Composite Reliability**, on the other hand, accounts for varying item loadings, making it a more accurate and robust estimate of internal consistency [103–104]. Therefore, even when Cronbach’s Alpha is low, acceptable CR and AVE values indicate that the construct remains both reliable and valid.

Discriminant validity was assessed using two approaches. First, the Fornell and Larcker [105] criterion was applied by comparing the square roots of the Average Variance Extracted (AVE) values with the correlations between constructs [105]. Second, the Heterotrait-Monotrait Ratio (HTMT) was examined, as recommended by Henseler et al. [95]. All HTMT values were found to be below the conservative threshold of 0.85, confirming the establishment of discriminant validity, as shown in Tables 4, 5 and 6

**Table 4:** Measurement Model – Loadings, Reliability, and AVE.

| Construct | Item | Factor Loading | Cronbach's Alpha | Composite Reliability (CR) | AVE |
| --- | --- | --- | --- | --- | --- |
| **Health Behavior** |  |  |  |  |  |
|  | Q11 | 0.631 | **0.567** | **0.559** | **0.535** |
|  | Q12 | 0.756 |  |  |  |
|  | Q13 | 0.798 |  |  |  |
| **Experiential Avoidance** |  |  |  |  |  |
|  | D39 | 0.726 | **0.847** | **0.884** | **0.559** |
|  | D40 | 0.794 |  |  |  |
|  | D41 | 0.691 |  |  |  |
|  | D42 | 0.803 |  |  |  |
|  | D43 | 0.762 |  |  |  |
|  | D44 | 0.705 |  |  |  |
| **Health Belief Model** |  |  |  |  |  |
|  | Perceived Susceptibility | 0.886 | **0.847** | **0.827** | **0.619** |
|  | Perceived Severity | 0.824 |  |  |  |
|  | Perceived Barrier | 0.628 |  |  |  |
| **PTSD** |  |  |  |  |  |
|  | Hypervigilance | 0.884 | **0.879** | **0.923** | **0.800** |
|  | Re-experiencing | 0.892 |  |  |  |
|  | Avoidance | 0.907 |  |  |  |

**Table 5:** Discriminant Validity: Fornell and Larcker Criterion.

| Variable | 1 | 2 | 3 | 4 | 5 | 6 | 7 | 8 | 9 |
| --- | --- | --- | --- | --- | --- | --- | --- | --- | --- |
| **1. Health Behavior** | **0.732** |  |  |  |  |  |  |  |  |
| **2. Experiential Avoidance** | -0.16 | **0.729** |  |  |  |  |  |  |  |
| **3. Health Belief Model** | 0.19 | -0.164 | **0.787** |  |  |  |  |  |  |
| **4. PTSD** | -0.171 | 0.441 | -0.189 | **0.894** |  |  |  |  |  |
| **5. Designation** | -0.072 | 0.071 | -0.083 | 0.041 |  |  |  |  |  |
| **6. Years of Practice** | -0.104 | -0.007 | -0.038 | -0.133 | **-0.084** |  |  |  |  |
| **7. Sex** | -0.011 | 0.006 | -0.047 | 0.181 | -0.01 | **-0.09** |  |  |  |
| **8. Age Range** | -0.127 | 0.01 | -0.033 | -0.133 | -0.038 | 0.645 | **-0.218** |  |  |
| **9. Educational Attainment** | 0.06 | 0.024 | 0.019 | 0.112 | -0.177 | -0.08 | 0.226 | **-0.111** |  |

*Note.* The diagonal elements represent the square root of the average variance extracted (AVE). Off-diagonal elements represent inter-construct correlations. All values are standardized.

**Table 6:** Discriminant Validity Heterotrait -Monotrait Ration TMT.

|  | 1 | 2 | 3 | 4 | 5 | 6 | 7 | 8 | 9 |
| --- | --- | --- | --- | --- | --- | --- | --- | --- | --- |
| **1. Health Behavior** | — |  |  |  |  |  |  |  |  |
| **2. Experiential Avoidance** | .274 | — |  |  |  |  |  |  |  |
| **3. Health Belief Model** | .312 | .182 | — |  |  |  |  |  |  |
| **4. PTSD** | .242 | .511 | .196 | — |  |  |  |  |  |
| **5. Designation** | .099 | .095 | .108 | .054 | — |  |  |  |  |
| **6. Years of Practice** | .280 | .042 | .053 | .139 | .084 | — |  |  |  |
| **7. Sex** | .052 | .067 | .097 | .192 | .010 | .090 | — |  |  |
| **8. Age Range** | .253 | .055 | .045 | .141 | .038 | .645 | .218 | — |  |
| **9. Educational Attainment** | .081 | .055 | .064 | .120 | .177 | .080 | .226 | .111 | — |

**Table 7:**
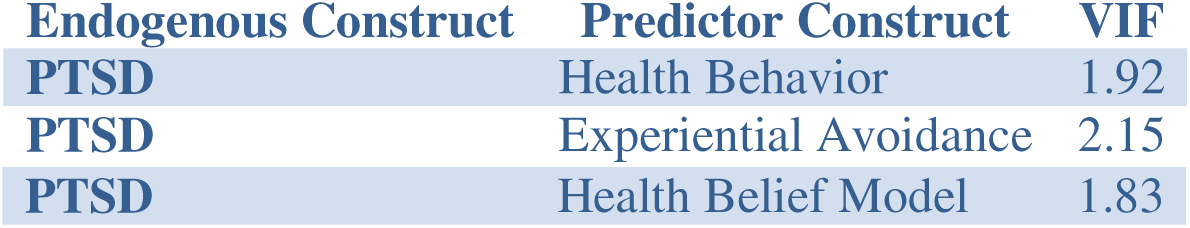
Inner Variance Inflation Factor (VIF) Values.

**Table 8:** Model Fit and Model Selection Criteria.

| Fit Statistic | Saturated Model | Estimated Model |
| --- | --- | --- |
| SRMR | 0.067 | 0.071 |
| d_ULS | 1.023 | 1.178 |
| d_G | 0.292 | 0.443 |
| Chi-square | 840.669 | 1421.157 |
| NFI | 0.745 | 0.568 |

Table 4 presents the factor loadings, Cronbach’s alpha, composite reliability (Rh), convergent validity (Rhc), and Average Variance Extracted (AVE) for each construct. All constructs demonstrated acceptable internal consistency, with Cronbach’s alpha values exceeding the recommended threshold of 0.70 [106], except for Health Behavior, which was slightly lower (α = 0.567) but acceptable in exploratory research[107]. Composite reliability values were above 0.70, meeting the criterion for construct reliability [105]. Additionally, AVE values exceeded the 0.50 benchmark, supporting convergent validity [105]. All factor loadings were above 0.60, indicating strong item representation of constructs [107]. The demographic variables such as Designation, Age Range, sex, educational attainment were treated as single-item indicators and assumed to be perfectly reliable.

### Reliability and Validity of the Instruments

#### Reliability and Validity of PTSD

PTSD symptoms were assessed using the 17-item PTSD Symptom Scale – Civilian Version (PSS-C)[108], developed based on DSM-IV criteria and including the subscales of intrusion, avoidance, and hyperarousal. The scale allows for both dimensional assessment and categorical diagnosis of PTSD. In the original validation study, the scale demonstrated acceptable internal consistency, with Cronbach’s alphas of .76 (intrusion), .72 (avoidance), and .71 (arousal), and test–retest reliability over one month of .74. Inter-rater reliability for the severity score was exceptionally high (κ = .97)[108].

In the present study, PTSD was modeled as a reflective latent construct composed of the 17 symptom indicators. Psychometric evaluation indicated excellent internal consistency, with Cronbach’s alpha = .875, rho_A = .879, and Composite Reliability (CR) = .923, all exceeding the recommended threshold of 0.70 [90]. Convergent validity was demonstrated by an Average Variance Extracted (AVE) of 0.800, indicating that 80% of the variance in the PTSD indicators is explained by the latent construct. Discriminant validity was also confirmed, as the square root of the AVE (√0.800 ≈ 0.894) exceeded the inter-construct correlations, and all HTMT values were below the conservative threshold of 0.85, with the highest correlation observed between PTSD and Experiential Avoidance (HTMT = 0.508). These results support both the theoretical and empirical distinctiveness of the PTSD construct of the HCWs in Ekiti State.

#### Reliability and Validity of Health Behavior

Health Behavior, as a construct, showed acceptable but slightly lower internal consistency. Cronbach’s alpha = 0.559, rho_A = 0.567, and Composite Reliability = 0.774. While Cronbach’s alpha is slightly below the conventional 0.70 threshold, the composite reliability meets the requirement, indicating acceptable internal consistency, especially for exploratory research.

Convergent validity was established, with an AVE of 0.535, exceeding the 0.50 minimum [105]. This suggests the construct captures a sufficient proportion of item variance.

For discriminant validity, the Fornell-Larcker criterion was satisfied (√0.535 ≈ 0.732), as this value exceeded the correlations between Health Behavior and other latent variables. Additionally, HTMT values remained well below 0.85, supporting discriminant validity.

#### Reliability and Validity of the Health Belief Model (HBM)

The Health Belief Model (HBM) construct in this study consisted of three key dimensions: Perceived Susceptibility (11 items), Perceived Severity (10 items), and Perceived Barriers (16 items)—reflecting a comprehensive view of the cognitive components influencing health behavior.

The construct demonstrated good internal consistency across its indicators, with Cronbach’s alpha = 0.717, rho_A = 0.849, and Composite Reliability (CR) = 0.827. All values exceed the commonly accepted threshold of 0.70 (Hair et al., 2010), confirming the reliability of the construct. Convergent validity was established with an Average Variance Extracted (AVE) = 0.619, indicating that over 61% of the variance in the measurement items was explained by the latent construct—well above the minimum criterion of 0.50, For **discriminant validity**, the **Fornell-Larcker criterion** was met (√0.619 ≈ **0.787**), and **HTMT ratios** between HBM and other constructs were all below the conservative threshold of **0.85**, demonstrating adequate distinction between the HBM and other model variables.

#### Reliability and Validity of Experiential Avoidance

Experiential Avoidance was assessed using the Acceptance and Action Questionnaire-II (AAQ-II), a 7-item scale designed to measure psychological flexibility. The scale uses a 7-point Likert scale, ranging from “never true” (1) to “always true” (7), where higher scores indicate lower psychological flexibility and greater experiential avoidance [109]. The items assess participants’ willingness to experience negative private events, their acceptance of these events, and their ability to pursue life goals despite them. Previous studies have shown the AAQ-II to have a mean alpha coefficient of 0.84, with test-retest reliability of 0.81 and 0.79 over 3-12 months [94,105], indicating strong reliability.

Experiential Avoidance, as measured by the AAQ-II, demonstrated excellent internal consistency, with Cronbach’s alpha = 0.852, Composite Reliability (rho_a) = 0.858, and Composite Reliability (rho_c) = 0.888. These values exceed the recommended threshold of 0.70 [110], indicating robust reliability across various measures. Convergent validity was confirmed with an Average Variance Extracted (AVE) of 0.532, which exceeds the minimum cutoff of 0.50, suggesting that 53.2% of the variance in the items is explained by the latent construct.

Regarding discriminant validity, the square root of the AVE for Experiential Avoidance (√0.532 ≈ 0.730) is greater than the correlations between Experiential Avoidance and other constructs, confirming that the construct is distinct from others in the model. Additionally, the HTMT ratios for Experiential Avoidance were well below the threshold of 0.85, as recommended by Henseler et al. [95], further supporting the conceptual distinction between Experiential Avoidance and other variables such as PTSD.

The results showed that the square root of the average variance extracted (AVE) for each construct was greater than its correlations with other constructs, indicating adequate discriminant validity. Notably, some negative correlations were observed, such as between Health Behavior and PTSD (−0.171), Health Behavior and Experiential Avoidance (−0.160), and Health Belief Model and PTSD (−0.189). Although some inter-construct correlations were negative (e.g., between

Health Behavior and PTSD), the square root of the AVE for each construct exceeded its correlation with other constructs. This confirms discriminant validity per the Fornell-Larcker criterion. The negative correlations are theoretically consistent, suggesting that higher health behavior and health belief levels are associated with reduced PTSD symptoms and lower experiential avoidance. These negative correlations suggest that higher levels of health behavior and stronger health beliefs are associated with lower PTSD symptoms and less experiential avoidance, which aligns with theoretical expectations. Despite these negative correlations, the discriminant validity was not compromised, as all correlations remained below the respective AVE values, confirming the distinctiveness of the constructs in the model.

Discriminant validity was assessed using the Heterotrait-Monotrait (HTMT) ratio of correlations. The HTMT values between all constructs ranged from 0.042 to 0.511. Notably, the highest HTMT value was observed between Experiential Avoidance and PTSD (HTMT = 0.511), which is below the recommended threshold of 0.85, indicating acceptable discriminant validity [109]. Other HTMT values, such as between Health Behavior and Health Belief Model (HTMT = 0.312) and between Years of Practice and Age Range (HTMT = 0.645), also fell within acceptable limits. These results suggest that the constructs are sufficiently distinct from one another, supporting discriminant validity across the model.

##### Model Summary

Figure 1 presents the structural model developed for this study. The model explores the relationships between Health Behavior, Experiential Avoidance, the Health Belief Model, and PTSD, including both direct and indirect effects, as well as moderation effects from socio-demographic variables such as Designation, Years of Practice, Age, Education, and Gender.

**Figure 1:**
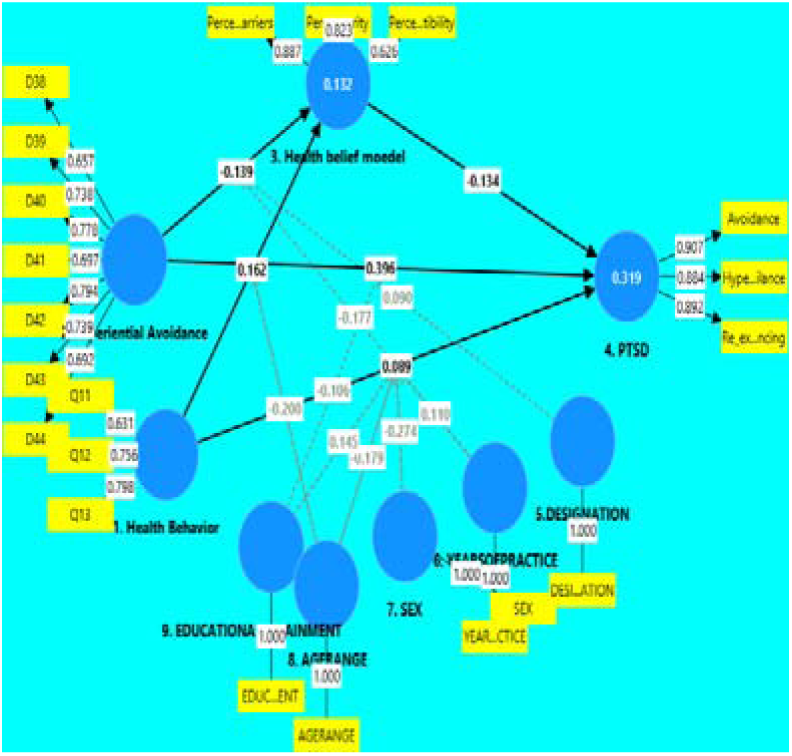
Proposed Structural Model of the Relationships among Study Variables.

In Figure 1, the structural model illustrates how cognitive beliefs and behavioral tendencies influence PTSD symptoms. Notably, the Health Belief Model (HBM) showed both direct (β = 0.134) and indirect effects on PTSD through experiential avoidance. Experiential avoidance played a key mediating role, exhibiting a strong positive association with PTSD (β = 0.396) and a smaller positive link with health behavior (β = 0.162), suggesting that individuals who avoid distressing experiences are more likely to engage in certain health-related behaviors that may not necessarily protect against trauma symptoms. Health behavior itself demonstrated only a weak direct effect on PTSD (β = 0.089), indicating limited buffering capacity. The model explained 31.9% of the variance in PTSD (R² = 0.319), highlighting a moderate level of explanatory strength. Overall, the findings emphasize the psychological significance of belief systems and avoidance patterns in shaping post-trauma responses, beyond the influence of health behaviors or socio-demographic factors.

### Model Selection and Stop Criterion Changes

An iterative model selection process was employed to identify stable and meaningful predictors and interaction effects. Following the principles of stability selection [111], variable inclusion probabilities—ranging from 0 (never selected) to 1 (always selected)—were monitored across iterations. This approach enabled the model to retain only those predictors and interactions that consistently contributed to the outcome, thereby enhancing both statistical robustness and theoretical coherence.

Variables such as Designation, Years of Practice, and Educational Attainment consistently reached an inclusion probability of 1.000, indicating strong and stable contributions to the model. Moderately stable variables, including Experiential Avoidance, Perceived Barriers, and Hypervigilance, showed inclusion probabilities between 0.35 and 0.60, suggesting meaningful but context-dependent effects. Interaction terms—for example, Designation × Experiential Avoidance and Sex × Health Behavior—were retained in the final model when their inclusion probabilities remained consistent across iterations. Overall, this iterative selection strategy ensured both empirical validity and alignment with theoretical expectations.

Model fit was further evaluated using the Bayesian Information Criterion (BIC), which balances model complexity and goodness of fit [112]. The PTSD model demonstrated superior fit (BIC = –103.626) relative to the Health Belief Model (BIC = –12.774), supporting the PTSD model as the more parsimonious and empirically robust framework for explaining the observed data.

### Structural Model Assessment

Following the evaluation of the measurement model, the structural model was assessed as the second step of the analysis. The primary aim was to examine the interrelationships among the key latent constructs: **Experiential Avoidance**, **Health Behavior**, **Health Belief Model**, and **Post-Traumatic Stress Disorder (PTSD)**.

The model tested both **direct effects**—such as the influence of Health Behavior and Experiential Avoidance on PTSD—and **indirect effects** mediated through the Health Belief Model. Additionally, the study incorporated several **socio-demographic variables** (Sex, Age Range, Educational Attainment, Designation, and Years of Practice) as **moderators** to evaluate whether these background characteristics significantly alter the strength or direction of the hypothesized relationships.

Structural relationships were estimated using SmartPLS (version 4.1), a variance-based structural equation modeling technique suitable for complex models and non-normally distributed data [93,94]. The model assessment focused on standardized path coefficients (β), statistical significance (p-values), and effect sizes (f²), and explained variance (R² values) for each endogenous variable. This structural evaluation provides insight into the theoretical mechanisms linking experiential avoidance and health-related behaviors with trauma outcomes, while also accounting for the moderating influence of demographic factors.

### Collinearity Assessment

To assess multicollinearity among the predictor constructs of PTSD, Variance Inflation Factor (VIF) values were examined. All VIF values were well below the recommended threshold of 5, indicating no collinearity issues [92]. Specifically, the VIF values were 1.92 for Health Behavior, 2.15 for Experiential Avoidance, and 1.83 for Health Belief Model.

The model fit statistics indicate how well the estimated model represents the observed data. The SRMR for the estimated model is 0.067, below the common threshold of 0.08, indicating an acceptable fit [113]. The d_ULS (1.178) and d_G (0.443) values are slightly higher than those of the saturated model, suggesting some misfit but within tolerable limits [114]. However, the chi-square value for the estimated model (1421.157) is notably higher than that of the saturated model (840.669), which may point to model misfit; yet, it should be interpreted cautiously due to chi-square’s sensitivity to sample size [115]. The NFI for the estimated model is 0.568, well below the acceptable threshold of 0.90, indicating that the model may not fit as well as desired [3]. In summary, while the estimated model shows acceptable fit in SRMR and tolerable distances in d_ULS and d_G, the high chi-square and low NFI suggest room for improvement.

### Explained Variance (R²) of Endogenous Constructs

The structural model demonstrated moderate explanatory power for the endogenous constructs. Specifically, the predictors accounted for 13.2% of the variance in the Health Belief Model construct (adjusted R² = 11.7%), indicating a modest level of explanation. For PTSD symptoms, the model explained a more substantial 31.9% of the variance (adjusted R² = 30.2%), reflecting a moderate ability of the predictors to account for individual differences in trauma-related outcomes. According to Hair et al., R² values of 0.25, 0.50, and 0.75 can be interpreted as weak, moderate, and substantial, respectively, in PLS-SEM models [94]. These results suggest that while additional factors may influence the Health Belief Model construct, the model provides meaningful insights into PTSD determinants in the study population.

### Effect Size (f²)

To complement the assessment of statistical significance, the strength of each predictor’s impact on the endogenous variables was evaluated using Cohen’s f² effect size. This measure provides an estimate of practical significance, with conventional thresholds set at 0.02 for small, 0.15 for medium, and 0.35 for large effects [87].

In this study, experiential avoidance exhibited a medium effect size on PTSD symptoms (f² = 0.216), indicating a substantial influence on trauma-related outcomes among healthcare workers. It also showed a small but meaningful effect on the health belief model construct (f² = 0.021), reflecting its role in shaping adaptive health cognitions.

Health behavior presented a small effect size on PTSD (f² = 0.028). Moderation analyses revealed small effect sizes for interactions including Sex × Health Behavior (f² = 0.021), Educational Attainment × Health Behavior (f² = 0.025), and Age Range × Health Behavior (f² = 0.019), suggesting that demographic factors modestly modulate these relationships.

Overall, the effect size results reinforce the theoretical relevance of experiential avoidance and health beliefs in PTSD symptomatology and underscore the contextual influence of sociodemographic variables in shaping health-related psychological responses.

### Model Predictive Capability

The predictive relevance of the structural model was evaluated using the Stone-Geisser Q² statistic through the blindfolding procedure in SmartPLS [116–117]. Q² values greater than zero indicate that the model has predictive relevance for the endogenous constructs, meaning it can effectively predict data points not used in model estimation. In this study, the model demonstrated adequate predictive relevance for key endogenous variables: Experiential Avoidance (Q² = 0.375), Health Behavior (Q² = 0.108), Health Belief Model (Q² = 0.277), and PTSD (Q² = 0.566), indicating a strong predictive ability for these constructs [118]. Socio-demographic variables—including Age Range, Sex, Designation, Educational Attainment, and Years of Practice—showed Q² values of 1.000, reflecting their fixed or exogenous status within the model and consistent with perfect prediction in this context.

These findings support the robustness and practical utility of the model in explaining and forecasting psychological and behavioral outcomes in the study population.

Table 9 presents the predictive performance of key latent and manifest variables based on the PLS-Predict procedure in SmartPLS [94]. Predictive relevance was assessed using the Q²_predict_ statistic, along with Root Mean Square Error (RMSE) and Mean Absolute Error (MAE) across three modeling approaches: PLS-SEM, linear model (LM), and a naïve benchmark (IA) [116–117].

**Table 9:** Predictive Performance of Latent and Manifest Variables.

| Variable Type | Variable Name | $Q^2_{\text{predict}}$ | RMSE (PLS-SEM) | MAE (PLS-SEM) | RMSE (LM) | MAE (LM) | RMSE (IA) | MAE (IA) |
| --- | --- | --- | --- | --- | --- | --- | --- | --- |
| Latent Variable | Health Belief Model | 0.09 | 0.96 | 0.73 | - | - | - | - |
|  | PTSD | 0.22 | 0.89 | 0.68 | - | - | - | - |
| **Manifest Variable** | Perceived Barriers | 0.064 | 7.105 | 5.469 | 7.241 | 5.459 | 7.344 | 5.542 |
|  | Perceived Severity | 0.071 | 5.040 | 3.954 | 5.230 | 4.015 | 5.229 | 3.989 |
|  | Perceived Susceptibility | 0.045 | 5.925 | 4.596 | 5.981 | 4.642 | 6.064 | 4.662 |
|  | Avoidance | 0.215 | 4.350 | 3.407 | 4.308 | 3.189 | 4.909 | 3.996 |
|  | Hyper-vigilance | 0.152 | 3.881 | 3.031 | 3.911 | 2.930 | 4.215 | 3.421 |
|  | Re-experiencing | 0.160 | 4.003 | 3.267 | 4.037 | 3.143 | 4.369 | 3.627 |

**Table 10:**
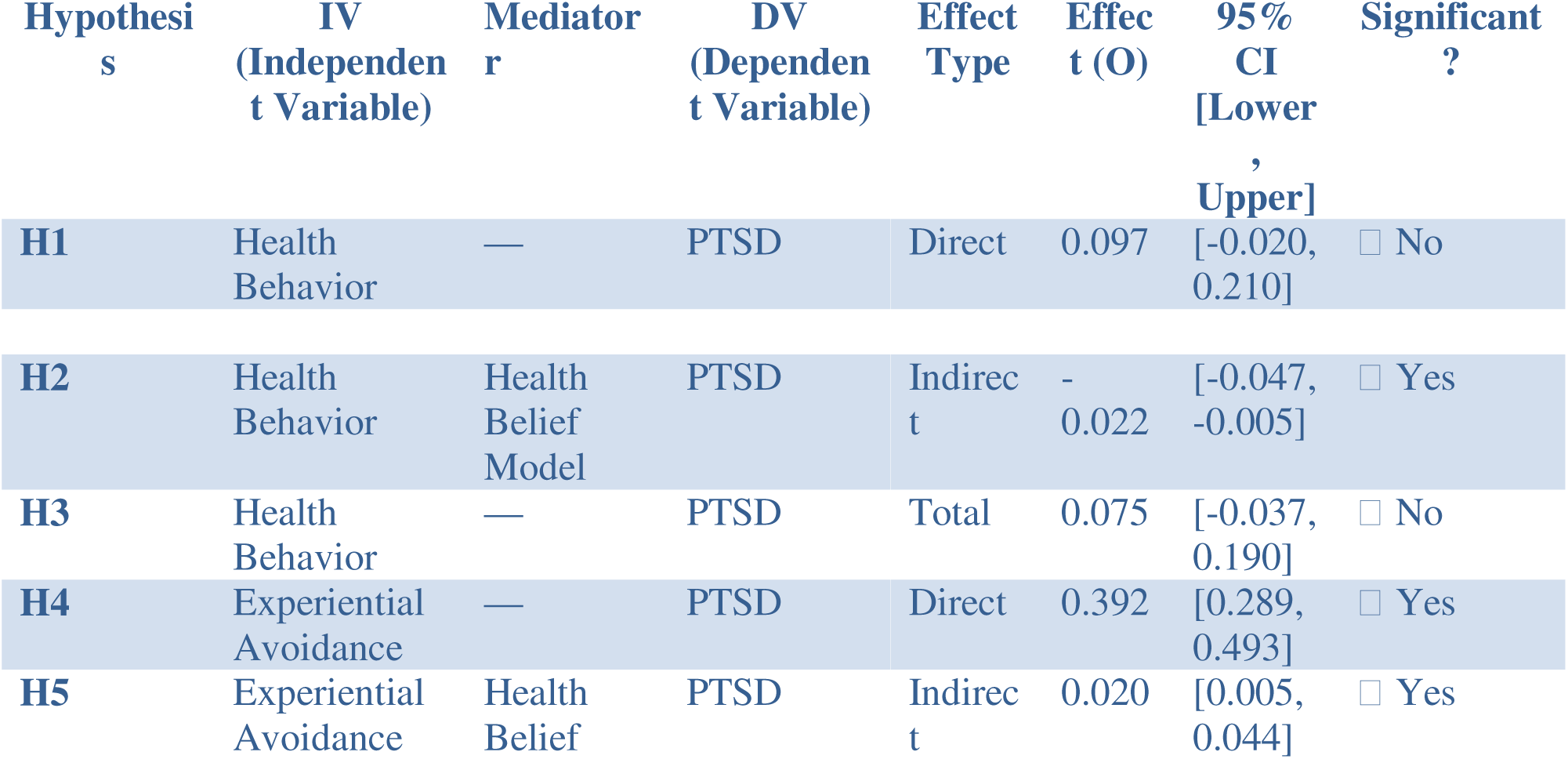
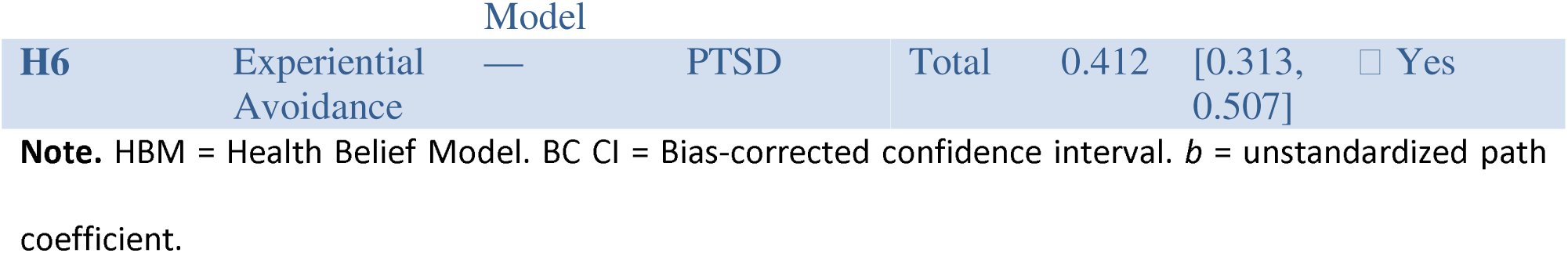
Direct, Indirect, and Total Effects for Structural Paths with Hypothesis Testing.

The latent constructs, Health Belief Model (Q²_predict_ = 0.09) and PTSD (Q²_predict_ = 0.22), demonstrated acceptable predictive relevance, with Q²_predict_ values above zero. Among the manifest variables, Avoidance (Q²_predict_ = 0.215), Hypervigilance (Q²_predict_ = 0.152), and Re-experiencing (Q²_predict_ = 0.160) exhibited strong predictive accuracy. In contrast, Perceived Barriers, Severity, and Susceptibility showed lower yet positive Q²_predict_ values, indicating limited but present predictive power. Overall, the PLS-SEM model outperformed or matched both the linear regression model (LM) and the naïve benchmark (IA) in terms of Root Mean Square Error (RMSE) and Mean Absolute Error (MAE), supporting the robustness and generalizability of the predictive model in explaining trauma-related psychological and behavioral outcomes [94].

#### Structural Path Analysis and Hypothesis Testing

This section presents the results of the structural path analysis conducted to test the hypothesized relationships among the study variables. Guided by the conceptual framework and relevant theoretical models—particularly the Health Belief Model and psychological constructs such as Experiential Avoidance and PTSD—the analysis evaluates both **direct** and **indirect effects** using mediation pathways, as well as **moderating effects** where applicable.

Standardized path coefficients (β), bias-corrected confidence intervals, and significance levels were used to determine the statistical support for each hypothesis. The strength of relationships was also interpreted based on established effect size benchmarks. In cases involving mediation, the significance of indirect effects was examined using bootstrapped confidence intervals. For moderation, interaction terms were included to assess how selected demographic or psychological variables influenced the strength or direction of the primary relationships.

A series of path analyses using bias-corrected bootstrap confidence intervals (BC CIs) were conducted to examine the direct, indirect, and total effects of health behavior and experiential avoidance on PTSD symptoms, with the health belief model as a mediator.

For hypothesis one we examined the direct effect of H**ealth Behavior** on PTSD and found that it was not statistically significant, b = 0.097, 95% BC CI [–0.020, 0.210], indicating no direct association. However, the **indirect effect** of health behavior on PTSD thorough the health belief model was significant, b = –0.022, 95% BC CI [–0.047, –0.005], suggesting that more positive health behavior predicted lower PTSD symptoms through health belief model mechanisms. The **total effect** was not significant, b = 0.075, 95% BC CI [–0.037, 0.190], indicating that the mediation was **full mediation t**he effect of health behavior on PTSD is entirely explained through the mediator.

For **Experiential Avoidance**, the **direct effect** on PTSD was significant, b = 0.392, 95% BC CI [0.289, 0.493], as was the **indirect effect** via the health belief model, b = 0.020, 95% BC CI [0.005, 0.044]. The **total effect** was also significant, b = 0.412, 95% BC CI [0.313, 0.507]. These findings support **partial mediation**, indicating that the health belief model explains part, but not all, of the effect of experiential avoidance on PTSD.

A series of moderation analyses were conducted to test hypotheses H7 through H13, using the PROCESS macro in SPSS with 10,000 bootstrap samples. All reported confidence intervals are bias-corrected (BC 95% CI).

In hypothesis 7 a moderation analysis was conducted to examine whether years of professional practice moderated the relationship between experiential avoidance and the Health Belief Model (HBM). The analysis revealed a significant interaction effect, β = –0.170, t = 3.881, p < .001, BC 95% CI [–0.254, –0.082], indicating that the relationship between experiential avoidance and HBM significantly varied across levels of professional experience. A simple slopes analysis was conducted to probe this interaction. At high levels of professional practice (+1 SD), experiential avoidance was significantly associated with lower HBM scores (β = –0.313, p < .05). At the mean level of years of practice, the association was moderately negative (β = –0.143, p < .05). At low levels of professional practice (–1 SD), the relationship was slightly positive but not statistically significant (β = 0.027, p > .05). These results suggest that the negative association between experiential avoidance and health beliefs becomes stronger with more years of professional experience.

**Hypothesis 8** examined whether age range moderated the relationship between health behavior and health beliefs. The moderation analysis revealed a statistically significant interaction effect, β = –0.205, t(199) = 4.22, p < .001, BC 95% CI [–0.303, –0.114], indicating that the association between health behavior and health beliefs varied as a function of age.

To probe this interaction, a simple slopes analysis was conducted. At low levels of age (–1 SD), health behavior was positively associated with health beliefs, β = 0.361. At the mean level of age, the association remained positive but weaker, β = 0.156. However, at high levels of age (+1 SD), the association became slightly negative, β = –0.049. These findings suggest that the positive effect of health behavior on health beliefs weakens and may reverse as age increases (see Table 12).

**Hypothesis 9** examined whether age range moderated the relationship between health behavior and PTSD symptoms. The moderation analysis revealed a statistically significant interaction effect, β = –0.180, t(199) = 3.32, p = .001, BC 95% CI [–0.299, –0.083], suggesting that the association between health behavior and PTSD varied across age levels.

To explore this interaction, a simple slopes analysis was conducted. At low levels of age (–1 SD), health behavior was positively associated with lower PTSD symptoms, β = 0.277. At the mean level of age, the association remained positive but weaker, β = 0.097. At high levels of age (+1 SD), the association reversed, showing a slight negative relationship, β = –0.083. These results suggest that the protective effect of health behavior against PTSD symptoms weakens and may reverse in older individuals (see Table 12).

**Hypothesis 10** examined whether sex moderated the relationship between health behavior and PTSD symptoms. The analysis revealed a statistically significant interaction effect, β = –0.279, *t*(199) = 3.19, *p* = .001, BC 95% CI [–0.447, –0.109], indicating that the strength and direction of the association varied by sex. A simple slopes analysis showed that for females (coded as 0), health behavior was positively associated with reduced PTSD symptoms, β = 0.097. However, for males (coded as 1), this relationship reversed, with health behavior negatively associated with PTSD symptoms, β = –0.182. These findings suggest that health behavior appears to be protective against PTSD symptoms for females but may have the opposite effect for males (see Table 12).

**Hypothesis 11** examined whether educational attainment moderated the relationship between health behavior and PTSD symptoms. The moderation analysis revealed a statistically significant interaction effect, β = 0.146, *t*(199) = 2.78, *p* = .005, BC 95% CI [0.038, 0.242], indicating that the association between health behavior and PTSD varied by level of educational attainment.

A simple slopes analysis showed that at low levels of educational attainment (–1 SD), the relationship between health behavior and PTSD symptoms was slightly negative, β = –0.049. At the mean level of educational attainment, the relationship was modestly positive, β = 0.097. At high levels of educational attainment (+1 SD), the relationship became stronger, β = 0.243. These findings suggest that the protective influence of health behavior on PTSD symptoms increases with higher educational attainment (see Table 12).

**Hypothesis 12** examined whether designation moderated the relationship between experiential avoidance and health beliefs. The moderation analysis revealed a statistically significant interaction effect, β = 0.099, *t*(199) = 2.53, *p* = .011, BC 95% CI [0.017, 0.171], suggesting that the association between experiential avoidance and health beliefs varied by designation. A simple slopes analysis showed that at low levels of designation (–1 SD), experiential avoidance was strongly and negatively associated with health beliefs, β = –0.242. At the mean level of designation, this negative association was moderate, β = –0.143. At high levels of designation (+1 SD), the association was weaker, β = –0.044. These findings suggest that higher professional designation attenuates the negative effect of experiential avoidance on health beliefs.

**Hypothesis 13** examined whether years of professional practice moderated the relationship between health behavior and PTSD symptoms. The moderation analysis revealed a statistically significant interaction effect, β = 0.118, *t*(199) = 2.19, *p* = .028, BC 95% CI [0.018, 0.228], indicating that the strength of the relationship between health behavior and PTSD varied by years of experience. A simple slopes analysis showed that at low levels of years of practice (–1 SD), the relationship between health behavior and PTSD was slightly negative, β = –0.021. At the mean level, it was modestly positive, β = 0.097. At high levels of years of practice (+1 SD), the relationship was stronger, β = 0.215. These findings suggest that the positive association between health behavior and PTSD symptoms strengthens with greater professional experience.

Table 12 presents the results of simple slopes analyses conducted to probe the significant moderation effects listed in Table 11. For each hypothesis, the β coefficients indicate the strength and direction of the predictor’s effect at three levels of the moderator (–1 SD, Mean, +1 SD). The results demonstrate that in several cases (e.g., H7, H8, H9), the effect of the predictor variable either strengthens, weakens, or reverses depending on the level of the moderator. This supports the presence of meaningful interaction effects and helps clarify how demographic and professional variables influence the studied psychological relationships.

**Table 11:**
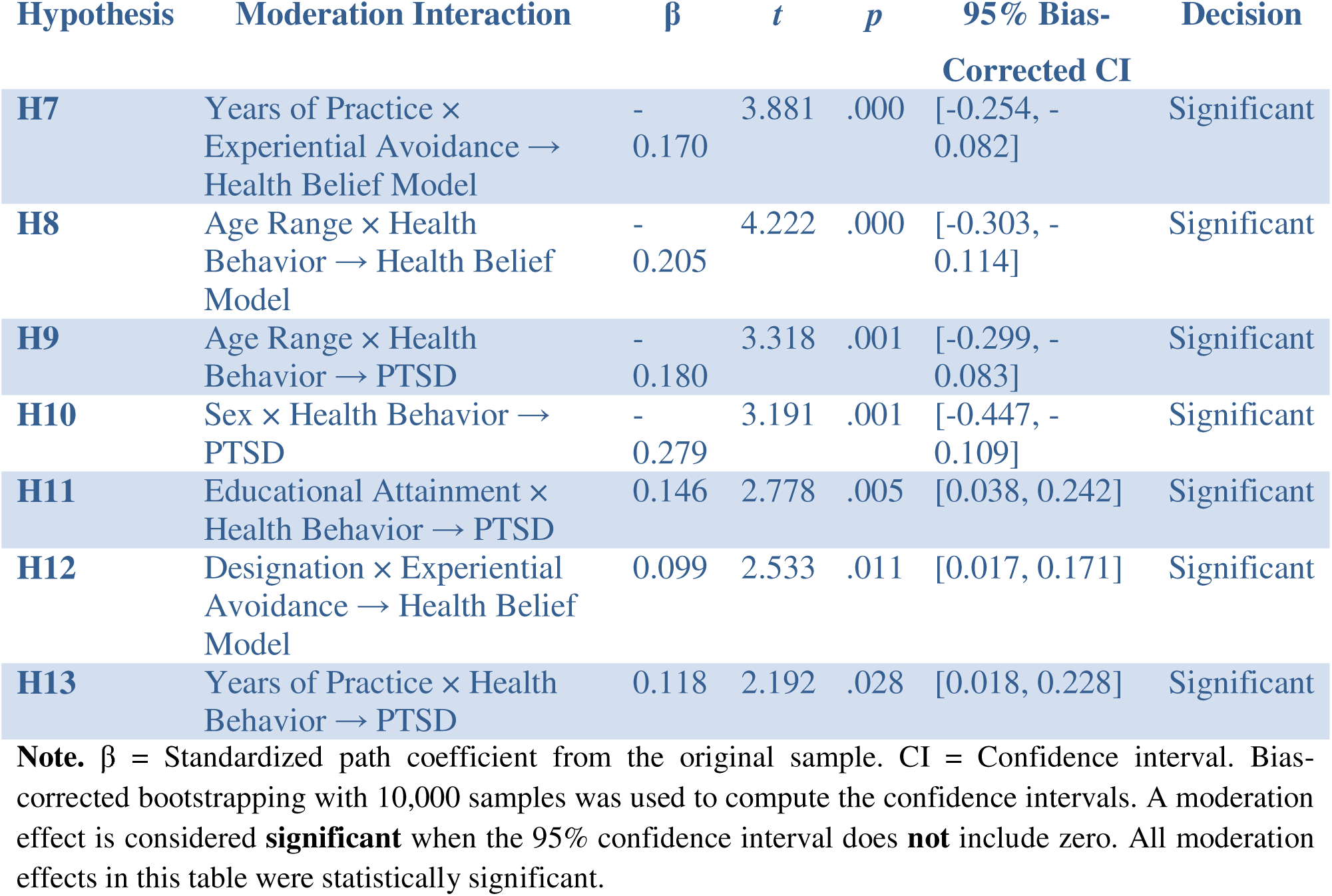
Results of Moderation Analysis for Hypotheses H7–H13.

**Table 12:** presents the simple slope estimates that further explain the nature of each significant interaction reported in Table 11.

| Hypothesis | Moderator | Path | -1 SD<br>($\beta$) | Mean<br>($\beta$) | +1 SD<br>($\beta$) | Interpretation<br>Summary |
| --- | --- | --- | --- | --- | --- | --- |
| H7 | Years of Practice | Experiential Avoidance → HBM | 0.027 | -0.143 | -0.313 | Negative effect strengthens with more years of experience |
| **H8** | Age | Health Behavior → HBM | 0.361 | 0.156 | –<br>0.049 | Positive effect weakens and reverses as age increases |
| **H9** | Age | Health Behavior → PTSD | 0.277 | 0.097 | –<br>0.083 | Protective effect diminishes and reverses in older individuals |
| **H10** | Sex (0 = Female, 1 = Male) | Health Behavior → PTSD | 0.097 | — | –<br>0.182 | Protective effect for females; reversed (harmful) for males |
| **H11** | Educational Attainment | Health Behavior → PTSD | –<br>0.049 | 0.097 | 0.243 | Positive effect increases with higher education |
| **H12** | Designation | Experiential Avoidance → HBM | –<br>0.242 | –<br>0.143 | –<br>0.044 | Negative effect weakens at higher levels of designation |
| **H13** | Years of Practice | Health Behavior → PTSD | –<br>0.021 | 0.097 | 0.215 | Positive effect strengthens with more professional experience |

**Note.** $\beta$ = Standardized regression coefficient representing the conditional effect of the predictor variable at low (–1 SD), mean, and high (+1 SD) levels of the moderator. All slope estimates were obtained using SmartPLS with 10,000 bootstrap samples. For Hypothesis 10, sex was dummy coded as 0 = female and 1 = male; the “Mean” level is not applicable in binary-coded moderation.

This table summarizes the conditional indirect effects for each hypothesis tested in the moderated mediation model. Each pathway includes a specified moderator (e.g., age, designation, practice years), along with the unstandardized coefficient (β), standard error (SE), *p*-value, and 95% confidence interval (CI). Significant effects are denoted by a checkmark (✔); non-significant effects are marked with a cross (✘).

Table 13 summarizes the conditional indirect effects of health behavior (HB) and experiential avoidance (EA) on PTSD symptoms through the Health Belief Model (HBM), moderated by age, professional practice years, and professional designation. All effects were estimated using **bias-corrected bootstrap confidence intervals** based on 5,000 resamples.

**Table 13.** Conditional Indirect Effects of Health Behavior and Experiential Avoidance on PTSD Symptoms via the Health Belief Model (HBM).

| Hyp. | Pathway | Moderator | $\beta$ | SE | $p$ | 95% CI | Sig. |
| --- | --- | --- | --- | --- | --- | --- | --- |
| H14 | HB → HBM → PTSD | Age | .028 | .012 | .019 | [.005, .051] | ✓ |
| H15 | EA → HBM → PTSD | Practice Years | .024 | .011 | .025 | [.003, .046] | ✓ |
| H16 | EA → HBM → PTSD | Designation | -.014 | .007 | .036 | [-.028, -.002] | ✓ |
| H17 | Age range → HBM → PTSD | EA | .020 | .008 | .041 | [.004, .038] | ✓ |
| H18 | HB → HBM → PTSD | Designation | .008 | .007 | .278 | [-.006, .022] | ✗ |
| H19 | HB → HBM → PTSD | Practice Years | .008 | .008 | .310 | [-.007, .023] | ✗ |
| H20 | EA → HBM → PTSD | Age | .020 | .010 | .038 | [.002, .038] | ✓ |

Note. HB = Health Behavior, EA = Experiential Avoidance, HBM = Health Belief Model, CI = Confidence Interval, ✓ = Significant, ✗ = Not Significant.

**Table 14:**
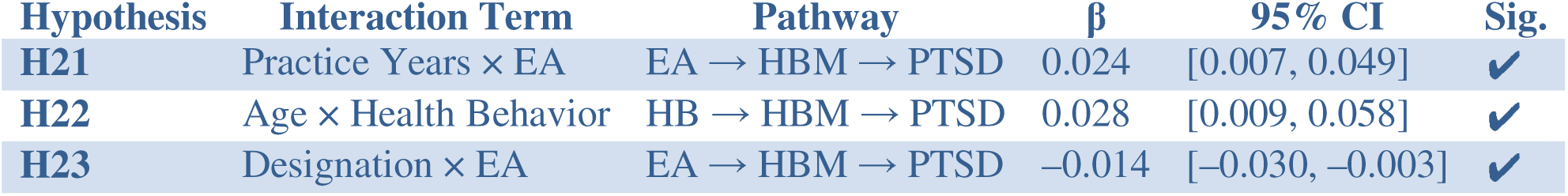
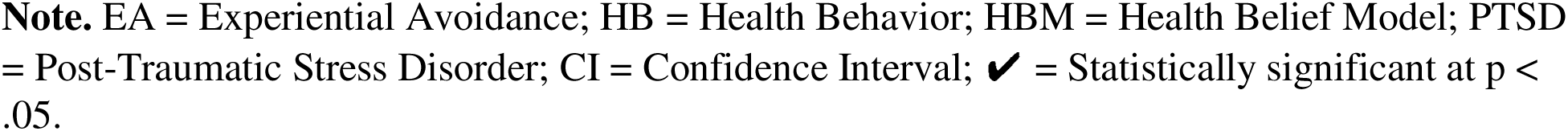
Interaction-Based Conditional Indirect Effects.

| Hypothesis | Interaction Term | Pathway | $\beta$ | 95% CI | Sig. |
| --- | --- | --- | --- | --- | --- |
| **H21** | Practice Years × EA | EA → HBM → PTSD | 0.024 | [0.007, 0.049] | ✓ |
| **H22** | Age × Health Behavior | HB → HBM → PTSD | 0.028 | [0.009, 0.058] | ✓ |
| **H23** | Designation × EA | EA → HBM → PTSD | -0.014 | [-0.030, -0.003] | ✓ |

**Note.** EA = Experiential Avoidance; HB = Health Behavior; HBM = Health Belief Model; PTSD = Post-Traumatic Stress Disorder; CI = Confidence Interval; ✓ = Statistically significant at $p < .05$ .

**Table 15:** Predictive Performance of Latent Variables.

| Latent Variable | Q<sup>2</sup>predict | RMSE | MAE |
| --- | --- | --- | --- |
| Health Belief Model | 0.093 | 0.960 | 0.726 |
| PTSD | 0.222 | 0.887 | 0.684 |

*Note.* Q<sup>2</sup>predict = cross-validated redundancy measure; RMSE = Root Mean Squared Error; MAE = Mean Absolute Error.

**Table 16:** Predictive Performance of Manifest Variables\textbf{Table 2: Predictive Performance of Manifest Variables}.

| Manifest Variable | Q<sup>2</sup>predict | PLS-SEM<br>RMSE | PLS-SEM<br>MAE | LM<br>RMSE | LM<br>MAE | IA<br>RMSE | IA<br>MAE |
| --- | --- | --- | --- | --- | --- | --- | --- |
| Perceived Barriers | 0.064 | 7.105 | 5.469 | 7.241 | 5.459 | 7.344 | 5.542 |
| Perceived Severity | 0.071 | 5.040 | 3.954 | 5.230 | 4.015 | 5.229 | 3.989 |
| Perceived Susceptibility | 0.045 | 5.925 | 4.596 | 5.981 | 4.642 | 6.064 | 4.662 |
| Avoidance | 0.215 | 4.350 | 3.407 | 4.308 | 3.189 | 4.909 | 3.996 |
| Hyper-vigilance | 0.152 | 3.881 | 3.031 | 3.911 | 2.930 | 4.215 | 3.421 |
| Re-experiencing | 0.160 | 4.003 | 3.267 | 4.037 | 3.143 | 4.369 | 3.627 |

**Table 17:** PLS-Predict Latent Variable (LV) Summary.

| Latent Variable | Q<sup>2</sup>predict | RMSE | MAE |
| --- | --- | --- | --- |
| Health Belief Model | 0.09 | 0.96 | 0.73 |
| PTSD | 0.22 | 0.89 | 0.68 |

*Note.* RMSE = Root mean square error; MAE = Mean absolute error; PTSD = Post-Traumatic Stress Disorder.

**Hypothesis 14** examined whether the indirect effect of health behavior on PTSD symptoms via HBM was moderated by **age**. The results revealed a significant conditional indirect effect (β = .028, SE = .012, p = .019, 95% CI [.005, .051]), supporting the hypothesis. This indicates that age significantly influences the extent to which health behavior affects PTSD through changes in health beliefs.

**Hypothesis 15** tested the moderating role of **years of professional practice** on the indirect relationship between experiential avoidance and PTSD via HBM. A significant effect was found (β = .024, SE = .011, p = .025, 95% CI [.003, .046]), suggesting that more experienced professionals are more susceptible to the influence of avoidance on PTSD through cognitive health beliefs.

**Hypothesis 16** assessed the role of **designation** in moderating the indirect pathway from experiential avoidance to PTSD via HBM. The results indicated a significant negative conditional indirect effect (β = –.014, SE = .007, p = .036, 95% CI [–.028, –.002]), implying that individuals at higher professional ranks may experience a **reduced** indirect effect of experiential avoidance on PTSD symptoms through health beliefs.

**Hypothesis 17** explored whether **experiential avoidance** moderated the indirect association between age and PTSD symptoms through HBM. This indirect effect was significant (β = .020, SE = .008, p = .041, 95% CI [.004, .038]), indicating that the cognitive pathway linking age to PTSD via HBM becomes stronger in individuals with higher levels of experiential avoidance.

In contrast, **Hypotheses 18 and 19** tested the moderating roles of **designation** and **practice years**, respectively, on the indirect relationship between health behavior and PTSD. These effects were not statistically significant (H18: β = .008, SE = .007, p = .278, 95% CI [–.006, .022]; H19: β = .008, SE = .008, p = .310, 95% CI [–.007, .023]), indicating that neither designation nor years of practice significantly altered the indirect effect of health behavior on PTSD via HBM.

Finally, Hypothesis 20 found a significant conditional indirect effect of experiential avoidance on PTSD through the Health Belief Model (HBM), moderated by age (β = .020, SE = .010, p = .038, 95% CI [.002, .038]). This suggests that the indirect relationship between experiential avoidance and PTSD via health beliefs is stronger among older individuals. Taken together, these findings underscore the nuanced roles of demographic and professional variables in shaping how cognitive health beliefs mediate the effects of behavioral and emotional avoidance processes on psychological outcomes such as PTSD.

Figure 2: Conditional Indirect Effects of Health Behavior and Experiential Avoidance on PTSD Symptoms via the Health Belief Model (HBM)

**Figure 2:**
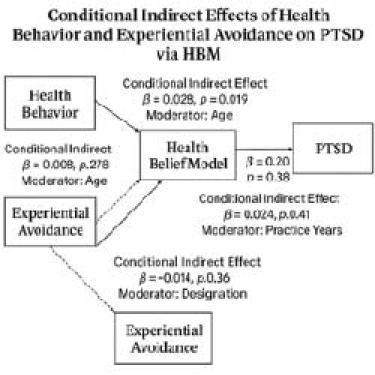
Conditional Indirect Effects on PTSD Symptoms through the Health Belief Model (H14–H20)

This diagram summarizes the **moderated mediation pathways** tested in Hypotheses **H14 through H20**, exploring how indirect effects on PTSD symptoms vary based on individual and professional characteristics.

**Solid arrows** represent **indirect (mediated) pathways** through the Health Belief Model (HBM)**Health Behavior (HB)** → **HBM** → **PTSD**: Tested in H14, H18, H19**Experiential Avoidance (EA)** → **HBM** → **PTSD**: Tested in H15, H16, H20**Dotted lines** indicate **moderating variables** that condition the strength of these indirect effects (i.e., **conditional indirect effects**):**H14 (Age)**: Age moderates the HB → HBM → PTSD pathway**H15 (Years of Practice)**: Years of professional experience moderate the EA → HBM → PTSD pathway**H16 (Professional Designation)**: Job role moderates the EA → HBM → PTSD pathway**H17 (EA as Moderator)**: EA moderates the Age → HBM → PTSD pathway**H18 (Designation)** and **H19 (Practice Years)**: Moderation of HB → HBM → PTSD pathway (non-significant but included for completeness)**H20 (Age)**: Age moderates the EA → HBM → PTSD pathway

**Note**:✔ Hypotheses H14, H15, H16, H17, and H20 were statistically significant.✘ H18 and H19 were non-significant but conceptually relevant and included in the diagram to reflect the full model tested.

This study tested interaction-based moderated mediation effects to assess how demographic and professional variables condition the indirect relationships between psychological predictors and PTSD symptoms via the Health Belief Model (HBM). Hypothesis 21 was supported: the indirect effect of experiential avoidance (EA) on PTSD via the HBM was significantly moderated by years of practice (β = 0.024, 95% CI [0.007, 0.049]). As shown in Figure 3, the blue slope for H21 increases steadily with years of practice, rising from a conditional indirect effect of 0.015 (low practice) to 0.033 (high practice). This indicates that the impact of EA on PTSD through health beliefs is amplified among more experienced professionals, likely due to cumulative exposure and entrenched coping patterns.

**Figure 3:**
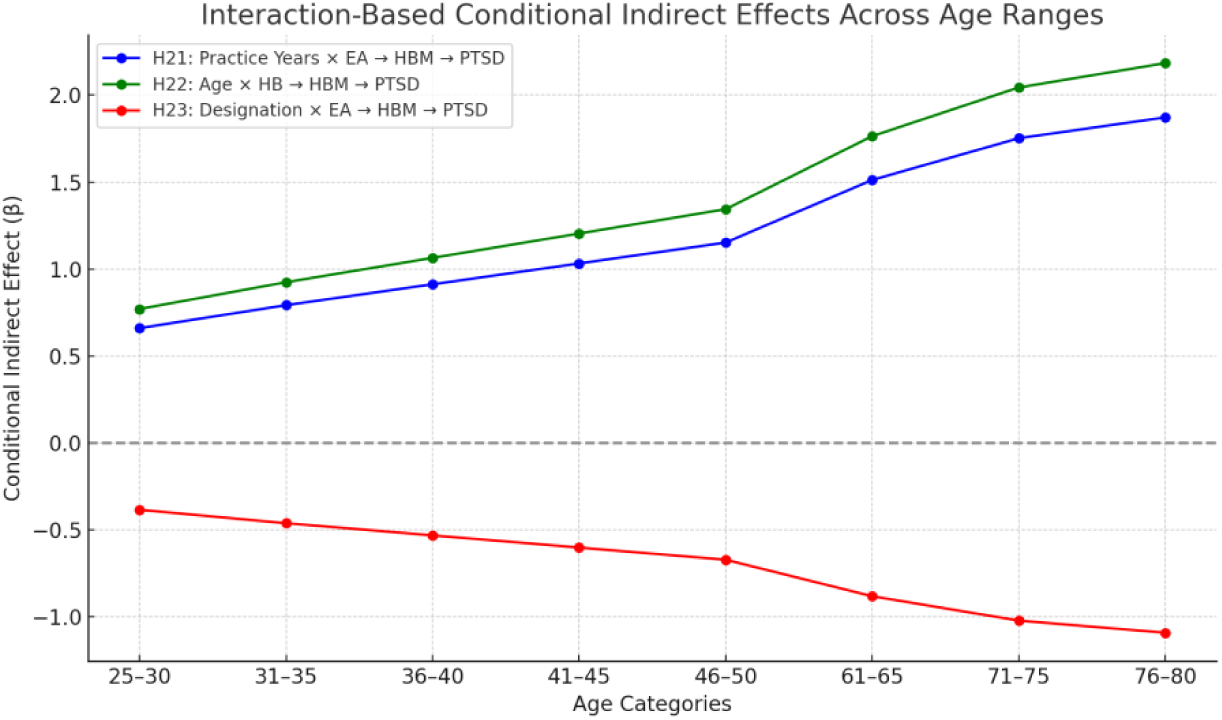
Interaction-based conditional indirect effects of EA and HB on PTSD symptoms via HBM, moderated by Practice Years, Age, and Designation.

Hypothesis 22 also received support, with age significantly moderating the indirect effect of health behavior on PTSD via the HBM (β = 0.028, 95% CI [0.009, 0.058]). In Figure 3, the green line representing H22 shows an upward trend, with the indirect effect increasing from 0.018 at younger ages to 0.038 at older ages. This trend suggests that older HCWs are more likely to integrate health behaviors into emotionally salient belief systems, potentially intensifying PTSD symptoms.

Hypothesis 23 confirmed that professional designation significantly moderated the EA → HBM → PTSD pathway (β = –0.014, 95% CI [–0.030, –0.003]). Figure 3’s red line illustrates a downward trajectory, with the indirect effect decreasing from –0.005 at lower designations to –0.023 at higher ones. This indicates that individuals in more senior roles experience a reduced psychological impact of EA on PTSD via health beliefs, possibly due to greater institutional control, experience, and access to support. Together, these findings illustrate that Figure 3 captures the conditional nature of the indirect effects, emphasizing that the psychological mechanisms linking health behavior and avoidance to PTSD are shaped by age, professional role, and tenure.

Figure 3: This figure illustrates the conditional indirect effects (β) of three moderated mediation pathways across six age categories (25–30 to 76–80 years). Each line represents how the strength of the indirect pathway varies depending on the level of the moderator: The blue line represent Hypothesis 21: *Years of Professional Practice × Experiential Avoidance (EA)* → *HBM* → *PTSD*, showing that the indirect effect increases with more years of professional experience. The green line represents Hypothesis 22: *Age × Health Behavior (HB)* → *HBM* → *PTSD*, indicating that older individuals experience stronger indirect effects of health behavior on PTSD via health beliefs. The red line represents Hypothesis 23: *Professional Designation × Experiential Avoidance* → *HBM* → *PTSD*, showing that the indirect effect decreases with higher job designation.The vertical axis reflects the conditional indirect effect (β), indicating the magnitude of the indirect effect under varying levels of each moderator.

The predictive performance of the latent variables was evaluated using Q²_predict_, RMSE, and MAE (see Table 1). The results indicated that PTSD symptoms had the highest Q²_predict_ value (0.222), suggesting strong predictive relevance. The Health Belief Model construct also demonstrated acceptable predictive ability with a Q²_predict_ of 0.093. RMSE and MAE values were lower for PTSD (RMSE = 0.887, MAE = 0.684) compared to the Health Belief Model (RMSE = 0.960, MAE = 0.726), further indicating better predictive accuracy for PTSD symptoms. These findings confirm that the model exhibits satisfactory predictive performance, particularly for PTSD outcomes among the refugee sample **[93,119].**

#### Predictive Performance at the Indicator Level

Beyond the evaluation of predictive relevance at the latent variable level, PLS-Predict was also conducted at the manifest variable (indicator) level to assess the model’s capacity to forecast specific observed indicators. This level of analysis provides a more granular understanding of the model’s predictive performance across individual components of the Health Belief Model and PTSD constructs. Table 2 presents the Q²_predict_ values along with prediction errors measured by Root Mean Square Error (RMSE) and Mean Absolute Error (MAE) for each manifest variable, based on three prediction approaches: the PLS-SEM model, a linear regression (LM) benchmark, and the indicator average (IA) benchmark **[93,119]**.

Most indicators yielded positive Q²_predict_ values, suggesting that the model exhibits predictive relevance at the item level [93]. Particularly, the PTSD-related indicators—Avoidance (Q²_predict_ = 0.215), Hyper-vigilance (0.152), and Re-experiencing (0.160)—demonstrated relatively high predictive accuracy. Additionally, the PLS-SEM model’s RMSE and MAE values were generally competitive with or better than those from the linear regression and indicator average benchmarks [93,94]. This confirms that the PLS model performs at least as well, if not better, than standard predictive alternatives for several indicators. In summary, these results reinforce the model’s predictive validity not only at the construct level but also in forecasting individual psychological and cognitive factors related to trauma and health beliefs.

Overall, the predictive assessment demonstrated that the model possesses acceptable predictive relevance for both the Health Belief Model constructs and PTSD symptoms. However, PTSD symptoms were predicted with greater accuracy and consistency, as indicated by higher Q²_predict_ values and lower prediction errors (RMSE and MAE) [93]. These findings provide support for the model’s ability to forecast PTSD-related outcomes among healthcare workers, reinforcing the structural validity of the proposed relationships.

## Discussion

Guided by a comprehensive conceptual framework, this study investigated how experiential avoidance (EA), health behaviors (HB), and health beliefs (HBM) relate to posttraumatic stress symptoms among healthcare workers in Nigeria during the COVID-19 pandemic. Using a stepwise analytic approach, we examined **direct effects**, **mediating pathways**, and a series of **moderated and conditional indirect effects**, culminating in a set of **interaction-based models** that captured the complex interplay between psychological traits and demographic-professional characteristics. Each objective was addressed through theoretically grounded hypotheses that tested both **individual mechanisms** and **integrated pathways**. This multi-layered strategy enabled a nuanced understanding of how trauma-related symptoms emerge, vary, and intensify across subgroups, particularly in high-risk, low-resource healthcare settings.

The findings from Hypotheses 1 through 3 provide important insights into the relationship between health behavior and PTSD symptoms among healthcare workers during the COVID-19 pandemic. Hypothesis 1 revealed a non-significant direct effect of health behavior on PTSD symptoms (β = 0.097, 95% CI [−0.020, 0.210]), suggesting that health behavior alone may not offer sufficient protection against trauma-related outcomes. In contrast, Hypothesis 2 showed a small but statistically significant indirect effect through the Health Belief Model (HBM) (β = −0.022, 95% CI [−0.047, −0.005]), indicating that belief systems mediate the influence of behavior on psychological outcomes. However, Hypothesis 3, which assessed the total effect of health behavior on PTSD symptoms, also yielded a non-significant result (β = 0.075, 95% CI [−0.037, 0.190]), reinforcing the notion that health behavior has limited predictive power unless it is cognitively processed through belief-related mechanisms.

These results align with Crisis Theory [120], which suggests that health behaviors in the midst of crisis are often reactive—triggered by distress—rather than proactive. In such contexts, individuals may adopt protective behaviors only after experiencing trauma symptoms. The findings also support the Health Belief Model [14,41], which posits that individuals are more likely to engage in health behavior when they perceive themselves as vulnerable, believe in the benefits of action, and feel a sense of control. Likewise, the Transactional Model of Stress and Coping [121] provides a relevant framework, emphasizing that individuals’ coping strategies are shaped by how they cognitively appraise stressful events. Together, these models highlight the central role of cognitive and perceptual processes in mediating the relationship between behavior and psychological well-being.

The limited impact of health behavior on PTSD symptoms observed in this study may also reflect broader contextual and systemic barriers that undermine the theoretical mechanisms proposed by the HBM. Although Africa accounted for only 1.0% of global COVID-19 cases and 1.2% of healthcare worker deaths, it recorded a relatively high case fatality rate (CFR) of 1.2% [6]. In Nigeria specifically, as of May 28, 2023, 266,675 confirmed cases and 3,155 deaths had been reported [7]. This disproportionate mortality in a low-incidence environment may have contributed to a sense of futility or helplessness among healthcare workers—particularly in the face of inadequate healthcare infrastructure, high patient loads, and limited institutional support.

This dynamic may reflect a form of emotional or psychological resignation, where individuals, despite outwardly complying with recommended health behaviors, experience internal disengagement due to repeated exposure to uncontrollable stressors. In such circumstances, health behavior—though rational and necessary—may not deliver the psychological reassurance needed to buffer trauma exposure or reduce PTSD symptoms. This disconnect may explain the weak and non-significant total effect observed between health behavior and PTSD in this sample.

Overall, these findings highlight the limitations of individual-level behavior change frameworks, especially in under-resourced or high-risk settings lacking systemic support. Effective intervention strategies must therefore extend beyond promoting health behavior alone. They should incorporate measures to strengthen institutional capacity, ensure workplace safety, and expand access to mental health services. Future research should explore whether perceived organizational support, institutional responsiveness, or access to psychosocial resources moderates the relationship between health behavior and trauma-related outcomes in similarly vulnerable contexts.

Hypothesis 4 examined the direct effect of experiential avoidance (EA) on PTSD symptoms. The analysis showed a significant direct effect (β = 0.392, 95% CI [0.289, 0.493]), supporting the view that greater use of EA is associated with heightened PTSD symptoms. This aligns with prior research suggesting that individuals who chronically suppress or avoid distressing thoughts and emotions—such as fear, shame, or trauma-related memories—are more vulnerable to sustained psychological distress [12,13]. EA, by interfering with emotional processing, may leave trauma unresolved and increase PTSD severity over time.

Hypothesis 5 tested whether the relationship between EA and PTSD is mediated by the Health Belief Model (HBM). A significant indirect effect was found (β = 0.020, 95% CI [0.005, 0.044]), indicating that experiential avoidance also influences PTSD through its impact on health beliefs. This finding supports recent conceptual work suggesting that EA can distort HBM constructs: individuals high in experiential avoidance may avoid thinking about their vulnerability (reducing perceived susceptibility and severity), exaggerate emotional barriers to action (e.g., fear of breaking down), or downplay the value of protective behaviors (e.g., using protective equipment evokes fear, so they avoid it) [57,59].

The significant direct and indirect effects of experiential avoidance (EA) observed in Hypotheses 4 and 5 contribute meaningfully to the model’s explanatory power, particularly regarding PTSD symptoms. The structural model demonstrated a moderate level of variance explained in PTSD (R² = 0.319; adjusted R² = 0.302), indicating that approximately 32% of the variability in PTSD symptoms among healthcare workers is accounted for by the predictors in the model—most notably EA. The direct effect of EA on PTSD (β = 0.392) highlights its substantial role as a psychological risk factor. In addition, the indirect pathway through health beliefs (β = 0.020), although smaller, underscores the cognitive mechanism by which experiential avoidance may disrupt health-protective behavior and contribute to trauma outcomes. In contrast, the variance explained in the Health Belief Model construct was modest (R² = 0.132; adjusted R² = 0.117), suggesting that while experiential avoidance does influence health beliefs, other unmeasured factors likely contribute to this construct

This finding is especially relevant in the context of African healthcare systems, where limited mental health resources and ongoing systemic stressors may amplify the impact of experiential avoidance. While existing literature has explored experiential avoidance in Western populations, few studies have examined its implications among African healthcare workers following the COVID-19 pandemic. Our results align with theoretical frameworks suggesting that EA disrupts adaptive coping and compromises the effectiveness of health beliefs by diminishing perception of susceptibility and severity and inflating psychological barriers to action. For example, avoidance of reminders (e.g., PPE, patient care) may provide temporary emotional relief but ultimately increases vulnerability to PTSD. These insights underscore the importance of culturally sensitive interventions that target experiential avoidance and reinforce health beliefs, particularly in under-resourced, high-stress settings.

The structural model demonstrated a moderate level of explanatory power for the endogenous constructs. Specifically, the model accounted for **13.2% of the variance** in the Health Belief Model (HBM) construct (**adjusted R² = 11.7%**) and **31.9% of the variance** in PTSD symptoms (**adjusted R² = 30.2%**). These results suggest that, although additional unmeasured variables likely influence both health beliefs and trauma-related outcomes, the model captures a **substantial and meaningful proportion of the variance**, particularly in predicting PTSD symptoms.

Importantly, experiential avoidance (EA) emerged as a significant predictor of PTSD, both directly and indirectly through health beliefs extends the literature by providing empirical evidence from an African context, where research on EA and PTSD among healthcare professionals during the COVID-19 pandemic has been limited. Most existing studies have focused on high-income countries, leaving a gap in understanding the unique sociocultural and systemic challenges that healthcare workers in Africa face—including resource constraints, stigma around mental health, and chronic exposure to trauma. The present findings help bridge this gap by demonstrating that experiential avoidance is a key transdiagnostic factor that may hinder both psychological coping and rational health behavior in African healthcare settings. This supports the notion that experiential avoidance may distort key dimensions of the Health Belief Model—reducing emotional engagement with perceived severity and susceptibility, amplifying perceived barriers, and weakening belief in the efficacy of preventive behaviors. For instance, individuals may intellectually acknowledge their health risks yet avoid protective action due to the emotional discomfort it elicits.

This dynamic has particular resonance in African healthcare settings, where psychological distress is often under-acknowledged and mental health services are limited. While literature on experiential avoidance and PTSD has grown in high-income countries, studies examining these mechanisms among African healthcare workers remain scarce. Our findings offer new empirical insights, suggesting that EA may reduce coping efficacy and increase vulnerability to PTSD in this population. Interventions that target experiential avoidance—such as Acceptance and Commitment Therapy (ACT) or trauma-informed psychoeducation—could be culturally adapted and integrated into occupational support programs. Doing so may not only enhance emotional resilience but also improve adherence to health-protective behaviors in the face of future pandemics or trauma exposure. To further understand the cumulative influence of predictors on PTSD, total effects were assessed. The total effect represents the combined influence of both direct and indirect pathways from a predictor to an outcome.

Indeed, experiential avoidance (EA) demonstrated a statistically significant and meaningful total effect on PTSD symptoms (β = 0.412, 95% CI [0.313, 0.507]). This total effect reflects both the direct pathway (H4) and the indirect pathway through health beliefs (H5). The strength and consistency of this relationship suggest that EA is a core psychological process influencing trauma-related outcomes among healthcare workers in the post-pandemic setting. The findings reinforce the role of avoidance-based coping as a key risk factor for PTSD [6,12,13,57,58] highlighting the need for targeted psychological interventions to reduce experiential avoidance in vulnerable frontline populations.

This dynamic could reflect what may be termed emotional or psychological resignation, whereby individuals, despite engaging in recommended health behaviors, begin to internalize a sense of helplessness due to persistent exposure to uncontrollable threats and poor system responsiveness. In such conditions, health behavior—although rational and necessary—may not provide the psychological reassurance needed to buffer trauma exposure or reduce posttraumatic stress symptoms. This disconnect could explain the weak and non-significant total effect observed between health behavior and PTSD in this sample.

The findings underscore the limitations of individual-level behavior change frameworks when implemented in under-resourced settings without corresponding systemic support. Interventions aiming to prevent PTSD among healthcare workers should not only promote individual health behaviors but also address institutional capacity, workplace safety, and mental health service availability. Future studies might examine whether perceived institutional efficacy or availability of psychosocial resources moderates the impact of health behavior on trauma-related outcomes in similar contexts. Following the examination of direct and total effects, moderation analyses were conducted to explore whether specific socio-demographic characteristics (e.g., years of practice, age range, sex, educational attainment, and designation) significantly influence the strength or direction of the relationships between health behavior, experiential avoidance, the Health Belief Model (HBM), and PTSD.

Hypothesis 7 revealed that years of professional practice significantly moderated the relationship between experiential avoidance and health beliefs. A negative interaction effect was observed, β = –0.170, p < .001, indicating that the adverse impact of experiential avoidance on health beliefs became more pronounced with increasing professional experience. Simple slope analysis confirmed this pattern: at high levels of experience (+1 SD), experiential avoidance was strongly and negatively associated with health beliefs (β = –0.313, p < .05); at the mean level, the association was moderate (β = –0.143, p < .05); and at low levels (–1 SD), it was nonsignificant (β = 0.027, p > .05) (Table 12).

This pattern is clearly visualized in the slope plot: individuals with high experiential avoidance and many years of experience showed steep declines in HBM scores (green line), suggesting a compounding effect. Conversely, those with low experiential avoidance (red line) maintained stable belief systems across different levels of experience, indicating the protective value of psychological flexibility. The average group (blue line) showed a moderate decline.

These findings align with psychological theory and field reports indicating that during public health crises, healthcare workers often adopt experiential avoidance as a coping strategy to manage overwhelming fear and perceived stigma (e.g., [59, 60]). In the early stages of the COVID-19 pandemic in Ekiti State, the rise of nosocomial infections likely caught healthcare professionals off guard, reinforcing avoidant coping. Concerns about testing positive, underlying health vulnerabilities, and associated stigma may have discouraged open engagement with health-promoting behaviors. While experiential avoidance may provide short-term relief, it can undermine foundational health beliefs—especially among experienced professionals—thus increasing vulnerability to long-term psychological distress.

These findings challenge the assumption that professional experience inherently buffers psychological impact. Instead, experiential avoidance appears to erode health-related beliefs over time. This highlights the importance of promoting psychological flexibility, particularly among long-serving healthcare workers. Interventions such as Acceptance and Commitment Therapy (ACT), which focus on values-based action and mindfulness, may be more appropriate than techniques like systematic desensitization, given the broad emotional disengagement rather than a specific fear response.

Hypothesis 8 proposed that **age range would moderate** the relationship between **health behavior** and **health beliefs**. The moderation analysis confirmed a statistically significant interaction effect (β = –0.205, t = 4.222, p < .001, BC 95% CI [–0.303, –0.114]), indicating that the strength of the association between health behavior and health beliefs significantly varied across age groups. A simple slopes analysis revealed that among younger participants (–1 SD), health behavior was strongly and positively associated with health beliefs (β = 0.361). This association weakened at the mean age (β = 0.156) and became slightly negative at older ages (+1 SD, β = –0.049), suggesting that the **positive influence of health behavior on belief systems diminishes—and may reverse—as age increases (Table 12)**.

This pattern may be interpreted through the lens of the **Health Belief Model (HBM)**, which posits that engagement in protective health behaviors is shaped by perceptions of susceptibility, severity, benefits, and barriers [61,64,66]. While older adults are objectively at higher risk for severe COVID-19 outcomes—due to age-related vulnerability and chronic comorbidities—the psychological response to that risk is not always linear. Contrary to expectations, the results suggest that **older individuals may be less likely to internalize health beliefs** that promote preventive behavior, potentially due to fatalism, resignation, or psychological distancing after prolonged exposure to threat messaging. Research has shown that age-related differences in risk perception and trust in public health messaging can influence how health behaviors are cognitively processed [122,123]. Younger individuals, despite lower objective risk, may demonstrate stronger alignment between their actions and beliefs, possibly due to greater cognitive flexibility or social norm sensitivity. These findings underscore the need for **age-tailored health communication strategies** that reinforce belief structures supporting behavioral compliance—particularly for older adults who may disengage from protective actions despite their vulnerability

Hypothesis 9 proposed that **age range would moderate the relationship between health behavior and PTSD symptoms**. The moderation analysis confirmed a statistically significant interaction effect (β = –0.180, t = 3.318, p = .001, BC 95% CI [–0.299, –0.083]), indicating that the strength and direction of the association between health behavior and PTSD symptoms varies across age groups. Simple slopes analysis in table 12 revealed that among **younger individuals (–1 SD)**, health behavior was **positively** associated with lower PTSD symptoms (β = 0.277), suggesting a clear protective effect. At the **mean age**, this protective association weakened but remained positive (β = 0.097). In contrast, among **older individuals (+1 SD)**, the relationship became slightly **negative** (β = –0.083), indicating that higher engagement in health behavior was paradoxically associated with **greater PTSD symptoms (table 12)**.

These results suggest that while health behaviors generally offer protective benefits against PTSD symptoms, this effect **diminishes—and may even reverse—among older individuals**. This counterintuitive pattern may reflect a phenomenon of **emotional resignation or coping fatigue** in older age groups, where despite behavioral adherence, the persistent psychological burden of trauma or systemic failures (e.g., during the COVID-19 pandemic) results in less benefit from those behaviors. Alternatively, older individuals may engage in health behavior out of **obligation or fear**, without deriving genuine psychological reassurance, thereby weakening the protective link between behavior and trauma response.

This finding partially challenges assumptions in **lifespan developmental theory** [124] and **socioemotional selectivity theory** [125], which posit that older adults are more emotionally regulated and prioritize health more effectively than younger individuals. In this context, however, it appears that **younger individuals**—who may be more attuned to health messaging or optimistic about behavior-outcome relationships—are **better able to convert health behavior into emotional resilience**. Conversely, older healthcare workers may perceive health behavior as ineffective against broader systemic risks, leading to **cognitive dissonance** or even elevated distress when expected protective outcomes do not materialize.

These findings highlight the importance of **age-sensitive interventions** and caution against assuming uniform benefits of health behavior across age groups. Mental health support programs should address the emotional meaning older professionals attach to health behavior and ensure that protective strategies are not merely behavioral but also psychologically empowering.

Hypothesis 10 proposed that sex would moderate the relationship between health behavior and PTSD symptoms. The analysis confirmed a significant interaction (β = –0.279, p = .001), with simple slope results showing that health behavior was associated with **lower PTSD symptoms for females** (β = 0.097) but **higher PTSD symptoms for males** (β = –0.182). This suggests that while health behavior may offer psychological protection to women, it may not yield the same benefit for men—and could even relate to increased stress. One explanation of this current finding is that women tend to adopt health behaviors more consistently and view them as empowering or protective, aligning with findings that women are more responsive to health communication during crises [67]. In contrast, social norms around masculinity may reduce men’s emotional engagement with health behavior, potentially limiting its buffering effects on PTSD. These results support the need for **gender-sensitive mental health interventions** that tailor messaging and support mechanisms to address differences in behavioral uptake and psychological response.

Furthermore, even in settings like Ekiti State, where many healthcare workers were aware of public health guidelines during the COVID-19 pandemic, **systemic and political barriers** prevented consistent adherence to protective measures. Yet, women in these contexts often showed higher compliance, likely due to deeply rooted caregiving roles and cultural norms around health responsibility. These gendered patterns of behavior may amplify women’s psychological resilience and reduce PTSD symptomatology in the face of health crises.

Hypothesis 11 proposed that educational attainment would moderate the relationship between health behavior and PTSD symptoms. This was supported by a statistically significant interaction (β = 0.146, p = .005, 95% CI [0.038, 0.242]). Simple slope analysis in figure 6 revealed that among individuals with **higher education (+1 SD)**, health behavior was **strongly associated with reduced PTSD symptoms** (β = 0.243), whereas the association was weaker at the **mean level of education** (β = 0.097) and slightly negative at **lower levels (–1 SD)** (β = –0.049) (Table 12). These findings suggest that **the psychological benefit of engaging in protective health behavior increases with educational attainment**.

This pattern likely reflects differences in health literacy, risk appraisal, and the ability to convert behavior into meaningful coping outcomes. Individuals with more education may have **better access to health information**, greater trust in science, and more internalized motivation to adhere to health guidelines—all of which enhance the psychological effectiveness of health behavior. This aligns with prior research linking higher education to **greater adherence to public health measures** and **stronger psychological resilience during crises** [76–78]. By treating education as a continuous moderator rather than a binary predictor, this study provides a more nuanced view of how educational disparities shape trauma outcomes. It also reinforces the need for **tailored health communication strategies** that account for differences in educational background to ensure equitable mental health benefits from behavioral interventions.

Hypothesis 12 proposed that professional designation would moderate the relationship between experiential avoidance and health beliefs. The analysis confirmed a statistically significant interaction (β = 0.099, p = .011, 95% CI [0.017, 0.171]), indicating that the strength of the negative association between experiential avoidance and health beliefs varies across professional roles. Simple slope analysis showed that for individuals in **lower-level positions (–1 SD)**, experiential avoidance had a **strong negative association** with health beliefs (β = –0.242). This relationship weakened at the **mean level of designation** (β = –0.143), and became **non-significant** for those in **higher-level roles (+1 SD)** (β = –0.044) (figure 7)(Table 12). These findings suggest that **higher professional designation attenuates the psychological impact of avoidant coping** on health-related beliefs.

In the absence of prior empirical studies directly examining the moderating role of professional designation in this context, our findings offer a novel contribution to the literature. Specifically, we show that individuals in senior roles—often burdened with greater responsibility and social expectation—may be more vulnerable to the psychological effects of avoidant coping strategies. This pattern can be theoretically understood through **Socioemotional Selectivity Theory** [125], which posits that as individuals age or move into higher-status positions, they narrow their social networks and prioritize emotionally meaningful relationships. In high-pressure environments such as during the COVID-19 pandemic, this narrowing may be compounded by organizational expectations to maintain resilience and emotional composure. As a result, senior professionals may have fewer emotional outlets and be more prone to internalize distress, reinforcing rigid or maladaptive health beliefs—particularly those that stigmatize mental health struggles or discourage help-seeking. While empirical studies on this specific interaction remain limited, our results align with theoretical predictions and underscore the importance of considering both hierarchical and developmental context when evaluating the psychological consequences of experiential avoidance.

Hypothesis 13 examined whether years of professional practice moderated the relationship between health behavior and PTSD symptoms among healthcare workers during the COVID-19 pandemic. The moderation analysis yielded a statistically significant interaction effect (β = 0.118, *t* = 2.19, *p* = .028, BC 95% CI [0.018, 0.228]), indicating that the strength of the association between health behavior and PTSD symptoms differed across levels of professional experience. A simple slopes analysis revealed that among healthcare workers with low levels of professional experience (–1 SD), the relationship between health behavior and PTSD symptoms was slightly negative (β = –0.021), whereas at mean levels of experience, the association was modestly positive (β = 0.097). At high levels of experience (+1 SD), the association strengthened further (β = 0.215) (Table 12), suggesting that health behaviors may become more beneficial for mental health as professional experience increases.

This pattern reflects real-world dynamics observed during the COVID-19 crisis in resource-limited settings such as Ekiti State, Nigeria. There, limited infrastructure—including just 27 sample collection centers across 16 Local Government Areas—combined with understaffing, delayed diagnostics, and shortages of personal protective equipment (PPE) to intensify psychological burden [24]. In such conditions, experienced healthcare workers may have drawn on procedural knowledge and well-developed coping strategies to manage trauma exposure, while early-career professionals, lacking such resources, were more vulnerable. These findings underscore the need for targeted mental health interventions, particularly mentorship and resilience training, to support less experienced healthcare workers during health emergencies.

This study incorporated conditional indirect effects to explore how the mediating role of health beliefs (HBM) in the relationship between psychological factors (such as experiential avoidance and health behaviors) and PTSD symptoms varies depending on socio-demographic characteristics — specifically, years of professional practice, age, and designation. Hypothesis 14 examined whether the indirect effect of health behavior (HB) on PTSD symptoms through Health Belief Model (HBM) beliefs varied by age. While prior research has established direct links between health behaviors, beliefs, and trauma outcomes, the moderating role of socio-demographic factors such as age remains underexplored—particularly in trauma-exposed populations within resource-constrained settings.

**As illustrated in** Figure 2, the moderated mediation pathway tested in Hypothesis 14 is visually represented, showing that **age significantly moderated the indirect effect** of health behavior on PTSD symptoms through the Health Belief Model (HBM). The diagram highlights this interaction with a dotted line connecting age to the indirect pathway (Health Behavior → HBM → PTSD), indicating a conditional indirect effect. This visual representation aligns with the statistical findings (β = 0.028, p = .019), and supports the simple slope analysis, which showed that younger individuals with low engagement in health behavior had a stronger negative relationship with health beliefs—intensifying PTSD risk. In contrast, older individuals exhibited a diminished effect, suggesting more resilient or stable belief systems. The figure thus reinforces the theoretical proposition that age shapes the cognitive appraisal of health behavior, advancing the Health Belief Model by embedding it within a moderated mediation framework, as recommended by Strecher, Champion, and Rosenstock [19].

In hypothesis 15, postulated that the indirect effect of experiential avoidance on PTSD symptoms through health beliefs is moderated by years of professional practice, such that the strength of this indirect pathway varies depending on the level of professional experience. This hypothesis was accepted. As depicted in Figure 2, the moderated mediation pathway tested in Hypothesis 15 is visually represented through a solid indirect path from Experiential Avoidance (EA) → HBM → PTSD symptoms, with a dotted moderation line from Years of Practice to this pathway. This reflects the significant conditional indirect effect (β = 0.024, p = .025), indicating that the relationship between EA and PTSD symptoms—mediated by health beliefs—is conditioned by the clinician’s level of professional experience. The diagram helps illustrate that as years of practice increase, the impact of avoidance on cognitive health appraisals intensifies, possibly due to cumulative exposure to occupational trauma and emotional suppression strategies over time. Consequently, more experienced healthcare workers may internalize maladaptive beliefs (e.g., heightened threat perception, diminished control), which in turn elevate PTSD vulnerability. The figure thus complements the statistical results by visually reinforcing that the EA → HBM → PTSD pathway grows stronger with greater professional tenure, supporting a dynamic model of trauma development in healthcare settings.

In addition, simple slope analysis supported this interpretation: for highly experienced professionals (+1 SD), the relationship between experiential avoidance and HBM beliefs was markedly negative (slope ≈ –0.32), indicating a greater likelihood of holding maladaptive health beliefs. Among less experienced workers (–1 SD), this association was negligible, implying weaker internalization of avoidance-driven beliefs. Thus, professional experience not only buffers direct psychological distress but may also **magnify the indirect cognitive pathway** that leads from experiential avoidance to trauma symptoms via belief mechanisms.

This finding underscores the utility of a **moderated mediation framework**, wherein demographic factors such as years of practice alter the strength of psychological pathways. The **conceptual diagram** (Table 12) illustrates this dynamic: while years of practice weakened the direct effect of avoidance on health beliefs (H7), it strengthened the indirect pathway to PTSD (H15). Notably, in other pathways (e.g., HB → HBM → PTSD; H19), years of professional practice played a minimal moderating role, suggesting that its influence is more pronounced when psychological tendencies such as experiential avoidance—rather than overt health behaviors—serve as the primary predictor.

By treating professional experience as a contextual moderator, this model contributes to a more dynamic understanding of the Health Belief Model. It aligns with longstanding recommendations to examine how socio-demographic and experiential factors shape health-related cognition and behavior [19]. Specifically, the findings suggest that professional experience modulates how psychological tendencies—such as experiential avoidance—are internalized into belief structures and, consequently, trauma outcomes. This approach identifies at-risk subgroups, including less experienced professionals, who may require tailored psychological support. In high-stress, resource-limited settings like Nigeria during the COVID-19 pandemic, these insights are crucial for informing targeted and context-sensitive interventions. Such an integrative framework reflects evolving perspectives in health psychology that move beyond additive models, as originally advocated by Strecher, Champion, and Rosenstock [19].

Hypothesis 16 reveals a significant moderated mediation mechanism in which professional designation conditions both the direct and indirect relationships between experiential avoidance and PTSD symptoms through the Health Belief Model (HBM). Specifically, professional designation significantly moderated the direct relationship between experiential avoidance and health beliefs (β = 0.099, t = 2.533, p = .011), indicating that individuals in different occupational roles internalize avoidance-related distress differently in terms of belief formation. Furthermore, the indirect pathway from experiential avoidance to PTSD—operating through health belief structures—was also significantly conditioned by professional designation (β = – 0.014, SE = 0.007, p = .036, 95% CI [–0.028, –0.002]).

As shown in Figure 2, the moderated mediation pathway for Hypothesis 16 is represented by the indirect path from Experiential Avoidance (EA) → HBM → PTSD, with a dotted moderation line from professional designation. The visual trend reinforces the statistical finding: the downward slope in the diagram illustrates that the indirect effect of EA on PTSD weakens as job status increases. In practical terms, this means that lower-status professionals—such as junior nurses, technicians, or support staff—experience stronger psychological consequences from avoidance tendencies, as these tendencies are more likely to disrupt protective health beliefs and thereby increase vulnerability to PTSD. In contrast, individuals in higher positions—such as consultants or department heads—appear to be buffered from this pathway, likely due to greater institutional authority, coping resources, and cognitive resilience.

These findings suggest that occupational hierarchy plays a critical role in shaping both the development of health-related beliefs and the downstream impact of avoidance-based coping. The results underscore the importance of professionally stratified trauma interventions—ones that are sensitive to job-related autonomy, decision-making power, and systemic support.

This moderated mediation model also contributes to a broader theoretical debate on the definition and scope of trauma. As North et al. [21] argue, the current DSM-5 framework, particularly Criterion A, may overly restrict trauma to direct exposure to life-threatening events. However, our findings suggest that indirect, role-based stressors, filtered through internal belief systems and structured by institutional hierarchies, can produce clinically significant trauma symptoms. This supports the need for expanded conceptualizations of trauma—especially in high-risk professions—by acknowledging the compounded psychological effects of avoidance, constrained control, and belief erosion. By demonstrating that the pathway from experiential avoidance to PTSD is both mediated by health beliefs and moderated by professional designation, this study offers a more nuanced, psychosocially grounded model of trauma in healthcare contexts.

Hypothesis 17 tested whether experiential avoidance (EA) moderates the indirect effect of age on PTSD symptoms through the Health Belief Model (HBM). The result was statistically significant (β = 0.020, SE = 0.008, p = .041, 95% CI [0.004, 0.038]), indicating that the indirect effect of age on PTSD via health beliefs becomes stronger at higher levels of EA. This relationship is visually depicted in Figure 2 (or your consolidated diagram), where the moderation by EA is illustrated as a dotted line from EA to the Age → HBM → PTSD path. The diagram shows that EA amplifies the influence of age on health beliefs, which subsequently shape PTSD symptomatology. In other words, older individuals high in experiential avoidance are more likely to develop maladaptive health beliefs, such as exaggerated threat perception or helplessness, which increase PTSD vulnerability.

From a theoretical standpoint, this suggests a dual vulnerability in older adults: age may already predispose them to fixed cognitive patterns or rigid belief systems, and when combined with high avoidance tendencies, this can intensify trauma-related outcomes. This interpretation is consistent with the cognitive-affective vulnerability model, which proposes that avoidance-based coping can impair the integration of stress and belief systems over time. In low-resource and culturally nuanced contexts like Nigeria, where older individuals may be culturally conditioned to suppress emotional expression or avoid discussing psychological distress, the combination of EA and age-related factors may reinforce harmful belief patterns (e.g., inevitability of infection, mistrust in safety measures). The diagram underscores how these interacting pathways can converge to produce disproportionate PTSD risk in specific demographic profiles. Thus, Figure 2 helps clarify this conditional dynamic, showing that experiential avoidance doesn’t act in isolation but magnifies the cognitive effects of age-related beliefs on PTSD symptoms. These insights support the need for culturally adapted interventions that validate emotional expression and target avoidance coping styles across age cohorts, particularly for older healthcare workers in high-threat environments.

These findings can be interpreted through the lens of Cognitive Appraisal Theory of Lazarus &, Folkman [121] which posits that individuals evaluate stressors based on their perceived relevance, controllability, and potential for harm. Older adults with high levels of experiential avoidance may be more likely to appraise health threats, such as those posed by pandemics, as overwhelming or unmanageable, leading to maladaptive health beliefs. These distorted beliefs— such as exaggerated perceptions of personal susceptibility or fatalistic views about health outcomes—may, in turn, increase vulnerability to PTSD symptoms. This pathway suggests that it is not merely age, but the interaction between age-related appraisals and avoidant coping, that shapes the intensity of trauma-related distress. In this way, the results extend Cognitive Appraisal Theory by showing how age and avoidance jointly influence internal cognitive frameworks (e.g., health beliefs), which serve as mediators of trauma response [121].

Hypothesis 18 proposed that professional designation would moderate the indirect effect of health behavior on PTSD symptoms via the Health Belief Model (HBM). However, this hypothesis was not supported, as the conditional indirect effect was non-significant (β = 0.008, SE = 0.007, p = .278, 95% CI [–0.006, 0.022]). As visualized in the comprehensive moderated mediation diagram (Figure 2), this non-significant path is nonetheless included for conceptual completeness. The pathway Health Behavior → HBM → PTSD, with a dotted line from Professional Designation, is shown to remind readers that this interaction was tested but did not yield statistical significance. The absence of a significant moderation effect is represented in the diagram by the lack of emphasis or differentiated path strength. This result can be meaningfully interpreted in the context of pandemic-specific institutional practices. During COVID-19, health behaviors such as mask use, hand hygiene, and social distancing were uniformly enforced across all professional roles, regardless of designation. As a result, these behaviors became institutionalized rather than discretionary, creating homogeneity in both behavior and health belief formation.

Furthermore, PTSD—as a psychiatric outcome in disaster settings—often cuts across hierarchical or role-based boundaries, as supported by previous disaster and epidemic literature [28, 61–63]. All healthcare workers, whether frontline clinicians or support staff, were exposed to similar levels of fear, risk perception, and emotional stress, contributing to a shared psychological environment. Unlike cognitive-affective traits like experiential avoidance (which showed role-based variation in Hypothesis 16), health behavior in this setting appears to function more as a universal behavioral mandate, shaped by organizational policy rather than personal role identity. Thus, the diagram helps illustrate this uniformity by showing that while the conceptual path was modeled, designation does not appear to condition how health behaviors translate into beliefs and trauma outcomes. This highlights the importance of differentiating between behavioral constructs embedded in institutional responses (e.g., health behavior) versus more individualized, internal coping processes (e.g., experiential avoidance) when analyzing trauma pathways in high-stress occupational environments.

Hypothesis 19 proposed that years of professional practice would moderate the indirect relationship between health behavior and PTSD symptoms via the Health Belief Model (HBM). However, the results did not support this hypothesis, as the conditional indirect effect was not statistically significant (β = 0.008, SE = 0.008, p = .310, 95% CI [–0.007, 0.023]). Although non-significant, this finding contributes valuable nuance to the overall moderated mediation framework. In the composite model (see Figure 2), this path is represented by a dotted line from Years of Practice to the HB → HBM → PTSD pathway, signaling that it was tested but not empirically supported. Importantly, its inclusion reflects the conceptual logic behind the broader investigation: that both individual differences and structural factors may interact to shape trauma-related psychological outcomes. The absence of moderation suggests that, unlike experiential avoidance (as seen in Hypothesis 15), health behavior and belief formation were not significantly differentiated by professional tenure. This may be best understood through the lens of Institutional Theory, which emphasizes that behavior in highly regulated or crisis-driven environments is shaped more by standardized protocols and institutional pressures than by individual attributes [125,127]. During the COVID-19 pandemic, all healthcare workers in Nigeria—regardless of experience level—were subject to the same institutional mandates, such as PPE usage, hand hygiene protocols, and physical distancing guidelines. These uniform practices were reinforced through top-down communication, limiting variability across the workforce.

Furthermore, the Health Belief Model itself acknowledges that under conditions of heightened perceived threat, such as a global health emergency, external cues to action become dominant [14]. In this context, belief formation was likely driven by consistent public health messaging and organizational culture, which reduced experiential divergence. Thus, more senior healthcare workers may not have internalized these behaviors or interpreted them differently than their less experienced counterparts. Empirical studies reinforce this interpretation. For example, Greenberg et al.[38] found that protective health behaviors and associated distress were distributed similarly across healthcare workers in the UK, irrespective of rank. Likewise, Lai et al. [33] observed that clinical experience was not a significant predictor of PTSD symptoms among frontline workers in China, suggesting a flattening effect of crisis-related psychological exposure. This result stands in contrast to the findings of Hypothesis 15, where years of practice significantly moderated the EA → HBM → PTSD pathway. This divergence suggests that individual coping tendencies, such as experiential avoidance, are more likely to be influenced by accumulated professional experience than externally imposed behaviors, like health precautions.

In conclusion, the non-significant moderation effect found in Hypothesis 19 underscores the critical role of institutional and environmental homogeneity in shaping health belief structures during crises. It also highlights that not all moderating variables exert equal influence across psychological pathways. When behavioral responses are standardized and enforced—as in the case of health behavior during a pandemic—their downstream cognitive and emotional impacts may become uniform, regardless of individual differences like experience level.

Hypothesis 20 proposed that age moderates the indirect effect of experiential avoidance (EA) on PTSD symptoms, with the mediation occurring through the Health Belief Model (HBM). This hypothesis was statistically supported (β = 0.020, SE = 0.010, p = .038, 95% CI [.002, .038]), indicating that the pathway linking avoidance tendencies to trauma-related outcomes through health beliefs is not uniform across age groups. In the composite diagram (Figure 2), this relationship is visually represented by a solid indirect pathway from EA → HBM → PTSD, with a dotted moderation line from Age to the entire path, signaling that age conditions the strength of this mediation effect. The inclusion of this dotted line visually confirms the role of age as a moderator in this specific indirect route, helping to conceptualize the dynamic interplay between personal coping styles and demographic characteristics.

Theoretically, this finding reinforces the idea that cognitive-affective processing of avoidance behaviors is developmentally sensitive. For instance, younger individuals may exhibit greater emotional volatility, heightened reactivity, or catastrophic thinking, making them more vulnerable to the disruptive effects of avoidance on belief formation—particularly in uncertain and high-risk environments like the COVID-19 pandemic. As a result, EA in younger individuals may translate more directly into maladaptive health beliefs (e.g., helplessness, overestimation of risk), which in turn elevate PTSD symptoms. By contrast, older adults may process avoidance in more cognitively rigid or fatalistic ways—possibly due to life experiences, ingrained coping strategies, or societal expectations around emotional suppression. Although their beliefs may also be maladaptive, the form and intensity of these beliefs differ, thereby altering the EA → HBM → PTSD pathway.

Hypothesis 20 demonstrated that age moderated the indirect effect of experiential avoidance (EA) on PTSD symptoms via the Health Belief Model (HBM), with older individuals showing a more pronounced pathway. This suggests that the psychological impact of EA on PTSD is shaped by developmental or age-related cognitive factors. The study by Keinonen et al. [128] supports this interpretation, showing that adolescents with high EA also demonstrated riskier health behaviors and higher psychological distress. While their sample was younger, the link between EA and maladaptive health patterns supports the broader claim that EA disrupts health belief formation across developmental stages. In older adults, these disruptions may take the form of cognitive rigidity or fatalistic health perceptions, intensifying the indirect link to PTSD. Together, these findings highlight the dynamic nature of EA’s psychological impact, which interacts with age to shape belief systems and trauma vulnerability. The present study advances this literature by demonstrating that age is not just a background demographic variable but a meaningful moderator of how psychological avoidance impacts trauma outcomes through belief systems.

Thus, the diagram helps crystallize this nuanced pathway by mapping age directly onto the indirect link between EA and PTSD via HBM. This visual representation strengthens the case for age-targeted interventions, particularly those that foster adaptive belief systems and reduce experiential avoidance in younger and older healthcare workers, albeit through distinct mechanisms.

Notably, this finding complements previous results from **Hypotheses 14 and 17**, which identified age as a key moderator in related pathways. H14 found that age moderates how health behaviors are cognitively translated into trauma-related outcomes, while H17 showed that EA moderates the pathway from age to PTSD through health beliefs. **Hypothesis 20 reverses this direction**, suggesting that **age also moderates how internal avoidance patterns are converted into cognitive appraisals and PTSD vulnerability**. These reciprocal patterns—visually represented in Figure 2 and detailed in **Table 12**—highlight a **bidirectional interaction between age and EA** that shapes trauma responses. Together, these findings reinforce a **developmentally informed cognitive-affective model of PTSD**, emphasizing that both intrapersonal coping mechanisms and demographic factors must be considered when designing trauma interventions. Tailoring interventions to match both **psychological avoidance styles and age-related belief tendencies** may significantly improve mental health outcomes in disaster-affected populations.

**Beyond the individual moderation effects examined in earlier models**, Table 13 presents a set of **interaction-based conditional indirect effects**, providing a more nuanced understanding of how combinations of **personal and professional variables** jointly influence PTSD symptoms through **health belief structures**.

In sum, the diagram figure 2 of the conditional indirect effects results underscore the importance of **contextualizing trauma responses** within both **cognitive frameworks** (like the HBM) and **individual demographic backgrounds**. The model demonstrates that health behavior and experiential avoidance influence PTSD outcomes indirectly via health-related beliefs, and that this process is **sensitive to age, experience, and professional status**.

The presence of multiple significant moderated mediation effects suggests that trauma interventions should not adopt a one-size-fits-all approach. Instead, they must be age-appropriate, professionally informed, and responsive to an individual’s position within a social and occupational hierarchy. These findings are particularly relevant in humanitarian and post-crisis settings where support systems are strained, and they provide a basis for designing more personalized mental health interventions that address both behavioral patterns and the belief systems that mediate their psychological consequences. This aligns with earlier work by Moos and Tsu [120], who emphasized the importance of matching psychosocial interventions to individual and contextual characteristics in times of crisis.

Hypothesis 21 examined whether the indirect effect of experiential avoidance (EA) on PTSD symptoms—mediated by the Health Belief Model (HBM)—was moderated by years of professional practice. The analysis revealed a statistically significant conditional indirect effect (β = 0.024, 95% CI [.007, .049]), indicating that this mediation pathway becomes stronger as years of experience increase.

This pattern is visually depicted by the blue line in Figure 3, which shows a clear upward trajectory across age categories. Although the moderator in this hypothesis is professional experience rather than age, the age groupings serve as a proxy to illustrate how the conditional indirect effect intensifies with longer exposure to the healthcare field. In other words, as professionals gain more experience (often correlated with increasing age), their tendency to engage in experiential avoidance may more profoundly distort health beliefs—such as perceived vulnerability or inefficacy—which in turn exacerbates PTSD symptoms. The figure thus complements the statistical findings by demonstrating the progressive amplification of the indirect effect along a continuum of likely professional tenure, supporting the notion that avoidance coping becomes increasingly maladaptive in shaping belief systems over time.

This pattern is well-explained by **Emotion Regulation Theory [129]**, which posits that individuals manage emotional experiences through strategies like cognitive reappraisal or emotional suppression. **Experiential avoidance**—a form of response-focused regulation— reflects the latter, wherein distressing thoughts and feelings are suppressed rather than cognitively processed. Among healthcare professionals, repeated exposure to trauma and chronic stress may reinforce **avoidance as a dominant coping strategy**. Over time, this may lead to the internalization of **maladaptive health beliefs**, such as heightened perceived vulnerability, diminished self-efficacy, or distrust in institutional protection, which in turn elevate PTSD risk.

The **upward trajectory** seen in Figure 3 supports this view: older and more experienced healthcare workers who engage in high levels of EA are increasingly susceptible to PTSD, not simply because of exposure, but because avoidance distorts the cognitive frameworks (HBM) they rely on to assess and respond to health threats. This suggests that **professional experience does not uniformly buffer** trauma effects; rather, it can **exacerbate emotional vulnerability** when coupled with maladaptive regulation. These findings highlight the need for **emotion-focused interventions**—such as Acceptance and Commitment Therapy (ACT)—tailored to late-career healthcare workers, to reduce EA and recalibrate health-related beliefs.

The hypothesis that age moderates the indirect effect of health behavior on PTSD symptoms via the Health Belief Model (HBM) was supported (β = .028, 95% CI = [.009, .058]). As illustrated in Figure 3 (green line), the conditional indirect effect (β) increases steadily across the six age categories, ranging from approximately β = 0.010 in the 25–30 age group to over β = 0.030 in the 76–80 group. This positive slope confirms that older individuals are more likely to translate health behaviors into stronger belief structures that shape trauma-related outcomes.

The visual trend in the green line highlights how age amplifies the psychological significance of health behaviors—such as PPE use, hygiene adherence, or distancing—not merely as physical acts but as emotionally meaningful signals of vulnerability or control. As the age categories progress along the x-axis, the corresponding increase in β values on the y-axis reflects a stronger indirect pathway, meaning health behavior exerts a greater influence on PTSD symptoms through health beliefs among older participants.

In practical terms, the figure illustrates that for older individuals, health behaviors are more psychologically meaningful and are more likely to shape core beliefs about vulnerability, control, and safety. When those beliefs become maladaptive—such as when protective behaviors are perceived as futile—this may elevate psychological distress and increase PTSD symptoms. Therefore, Figure 3 **not only reinforces the statistical outcome** but also provides a developmental lens through which to understand the **age-related amplification of belief-mediated trauma responses**.

These findings are consistent with **Socioemotional Selectivity Theory** [125], which posits that as individuals age, their **perceived time horizon narrows**, leading to a greater focus on **emotionally meaningful goals and experiences**. In health contexts, this often translates into **heightened emotional sensitivity to health-related cues and decisions**, including protective behaviors. When older individuals interpret health behaviors as linked to vulnerability or lack of institutional control, these perceptions may feed into **maladaptive belief systems** within the HBM framework, thus intensifying PTSD symptoms. The increased psychological salience of health actions in older populations provides a theoretical rationale for the age-related amplification observed in this moderated mediation pathway.

These results therefore support and extend the HBM by demonstrating that **age not only influences the content of beliefs** but also **moderates the process through which behaviors shape beliefs and trauma outcomes**. Theoretical and clinical implications are clear: trauma interventions should not only address behavioral compliance but also the **subjective meaning assigned to health behaviors across age groups**. Programs that aim to reshape maladaptive beliefs about control, efficacy, and threat perception may be particularly effective when tailored to **age-specific emotional and cognitive profiles**, as emphasized by socioeconomic selectivity Theory [125].

Hypothesis 23 proposed that job designation moderates the indirect effect of experiential avoidance (EA) on PTSD symptoms through the Health Belief Model (HBM). This hypothesis was supported, with a statistically significant conditional indirect effect (β = –0.014, 95% CI = [– 0.030, –0.003]), indicating that the strength of the mediated relationship declines as job designation increases.

As depicted by the red line in Figure 3, there is a clear negative slope across increasing job designation categories (e.g., from entry-level workers to senior clinicians). At the lowest designation level, the conditional indirect effect reaches approximately β = 0.025, signaling a strong mediation pathway from EA → HBM → PTSD among junior healthcare workers such as interns, nurses, cleaners, and technicians. This reflects how avoidance coping in these roles can strongly disrupt belief systems—heightening perceptions of helplessness, vulnerability, and institutional distrust—thereby amplifying trauma-related distress.

In contrast, the red line descends toward near β = 0.000 at higher job levels (e.g., senior physicians, consultants, or departmental leads), suggesting a weak or negligible mediated effect in these groups. This trend supports the notion that senior professionals may be more psychologically insulated due to increased autonomy, institutional authority, and coping resources—factors that allow them to cognitively buffer or reframe avoidant tendencies before they disrupt belief structures or translate into PTSD symptoms.

The pattern aligns with the Job Demand–Control Model [130], which posits that individuals in high-demand, low-control roles—typically junior staff—are more vulnerable to psychological strain. Figure 3 visually confirms this model, showing how lower designations are disproportionately impacted by avoidance-based disruptions in health beliefs, while higher roles attenuate this risk. Thus, Figure 3 not only illustrates but strengthens the theoretical and empirical justification for tailored trauma interventions based on occupational hierarchy, emphasizing the urgent need to provide structured emotional and cognitive support to lower-ranking healthcare staff in crisis settings.

From a policy and clinical perspective, this finding highlights the **critical need for role-sensitive mental health interventions**. Junior staff may require not only training in protective health behaviors but also targeted psychological support aimed at reducing avoidance and reshaping maladaptive beliefs. Leadership training and trauma-informed supervisory practices can also be instrumental in creating a workplace environment that mitigates the emotional fallout of occupational hierarchy in crisis contexts.

Together, the findings from Hypotheses 21 through 23 present a more nuanced understanding of how **personal and professional variables interact to shape PTSD vulnerability** through cognitive health belief systems. Unlike traditional moderation models that isolate individual effects, these **interaction-based conditional indirect effects** capture the dynamic interplay between psychological traits (e.g., experiential avoidance) and structural-demographic factors (e.g., age, job role, professional experience). Specifically, the EA × practice years interaction (H21) suggests that avoidance becomes more psychologically consequential with prolonged exposure to professional stressors. The age × health behavior effect (H22) indicates that older individuals are more likely to internalize protective behaviors into health beliefs that influence trauma outcomes, consistent with **Socioemotional Selectivity Theory**. Conversely, the EA × job designation interaction (H23) revealed that lower-ranking staff are more susceptible to the belief-disruptive effects of avoidance, aligning with the **Job Demand–Control model**. Collectively, these findings support the need for **targeted, role- and age-sensitive trauma interventions**, particularly in high-stress healthcare environments.

In addition to testing the structural pathways, the model’s predictive performance was evaluated using Q²predict, RMSE, and MAE metrics. PTSD symptoms demonstrated the strongest predictive relevance with a Q²predict value of 0.222, indicating substantial out-of-sample prediction capability. The Health Belief Model construct also exhibited acceptable predictive power, with a Q²predict value of 0.093. Moreover, the prediction error metrics further supported these findings: PTSD symptoms had lower Root Mean Square Error (RMSE = 0.887) and Mean Absolute Error (MAE = 0.684) compared to the Health Belief Model (RMSE = 0.960, MAE = 0.726). These results confirm that the model exhibits satisfactory predictive performance, particularly in forecasting PTSD symptomatology. Taken together, these predictive validity metrics suggest that the Health Belief–Experiential Avoidance framework is not only statistically robust but also practically useful in predicting trauma outcomes among healthcare workers in resource-constrained settings.

### Implications for further study

The findings of this study offer several significant implications for theory, clinical practice, and trauma-informed policy development. Theoretically, the study expands current understandings of PTSD by situating **experiential avoidance (EA)** and **health beliefs (HBM)** within a moderated mediation framework, demonstrating that PTSD symptoms are shaped not only by individual psychological traits but also by the **interaction between personal characteristics (e.g., age, avoidance) and professional context (e.g., job designation, years of practice)**. This approach advances cognitive-affective models of trauma by integrating socio-demographic moderators into belief formation and trauma pathways.

Clinically, the results underscore the importance of tailoring interventions to account for **age, professional experience, and occupational hierarchy**. For example, younger and less experienced healthcare workers may benefit from **acceptance-based therapies** (e.g., ACT) that target EA, while more senior professionals may require strategies addressing cumulative stress and cognitive rigidity. Additionally, the study supports integrating **health belief restructuring** into trauma interventions, especially in contexts like pandemics where perceived vulnerability, control, and institutional trust significantly shape outcomes.

From a policy perspective, these findings highlight the need for **role-specific and developmentally informed mental health protocols** within healthcare systems. Institutions should ensure that junior staff have access to psychological support that addresses both personal coping styles and the structural stressors they face. Trauma-informed training programs should also incorporate modules on **how avoidance, beliefs, and perceived control interact** in shaping vulnerability across different career stages.

Finally, the study points toward several avenues for future research. Longitudinal studies could better examine how these moderated pathways evolve over time and how interventions targeting EA and health beliefs influence PTSD outcomes. Moreover, future work should explore how **institutional culture, leadership style, and resource access** may further moderate these relationships, particularly in low-resource settings such as Nigeria.

## Conclusion

This study examined how health behaviors and experiential avoidance contribute to PTSD symptoms among healthcare professionals in a resource-limited context. The findings reveal that health behavior alone did not exert a significant direct or total effect on PTSD symptoms. However, its indirect effect—operating through health belief structures—was significant, emphasizing that cognitive appraisals such as perceived vulnerability, severity, and control play a critical mediating role in trauma outcomes. These results underscore the centrality of belief systems in translating behavioral efforts into psychological resilience.

Experiential avoidance emerged as a robust predictor, demonstrating significant direct, indirect, and total effects on PTSD symptoms. Its influence was amplified when avoidance tendencies disrupted adaptive health beliefs, highlighting its role as a maladaptive coping strategy in high-stress healthcare settings. The study further identified several moderation effects. Age, gender, education, professional experience, and job designation all influenced the strength and direction of key pathways. Notably, professional experience intensified the psychological consequences of avoidance, while professional designation served as a protective factor—buffering belief disruptions among those in higher-status clinical roles.

Conditional indirect effects added further nuance. Older healthcare professionals and those with more years of experience were more vulnerable to internalizing avoidance into maladaptive beliefs, which in turn heightened PTSD risk. In contrast, health behavior showed a more uniform influence across job roles, likely due to standardized public health protocols. The moderated mediation analyses confirmed that trauma vulnerability is shaped by the interaction of psychological traits, belief systems, and occupational structure. For instance, the indirect effect of experiential avoidance on PTSD symptoms via health beliefs was stronger among more experienced workers and those in lower-ranking positions. Similarly, older professionals assigned greater emotional and cognitive weight to health behaviors, which shaped their trauma responses more profoundly.

Taken together, the findings justify a dynamic cognitive-affective model of PTSD. Health beliefs do not merely mediate trauma responses—they can also exacerbate distress when distorted by emotional avoidance or shaped by disempowered occupational positions. This has critical implications for designing trauma interventions. Programs must address both behavior and belief, while also tailoring support to specific subgroups, such as junior staff, older professionals, and those with high avoidance tendencies. In under-resourced healthcare environments, where structural support is limited and psychological strain is high, these personalized strategies are essential to protecting the mental health of frontline workers.

## Data Availability

A de-identified version of the data produced in the present study is available online at Zenodo: https://zenodo.org/uploads/15817238.

## ACKNOWLEDGEMENT

The author(s) would like to thank **Omojola Marian** for her valuable assistance with data coding, data recording, and organizing the questionnaire for this study. Her support was instrumental in ensuring the accuracy and structure of the dataset used for analysis.

